# Composition and Function of the Gut Microbiome in Microscopic Colitis

**DOI:** 10.1101/2024.10.28.24316293

**Authors:** Albert Sheng-Yin Chen, Hanseul Kim, Etienne Nzabarushimana, Jiaxian Shen, Katherine Williams, Jenny Gurung, Jessica McGoldrick, Kristin E Burke, Long H. Nguyen, Kyle Staller, Daniel C Chung, Ramnik J Xavier, Hamed Khalili

## Abstract

**Background:** Microscopic colitis (MC) is a common cause of chronic diarrhea, predominantly among older adults. Emerging evidence suggests that perturbations of gut microbiome and metabolome may play an important role in MC pathogenesis.

**Objective:** To comprehensively characterize alterations of the gut microbial and metabolic composition in MC.

**Design:** We established a longitudinal cohort of adult patients with MC and two control groups of individuals – chronic diarrhea controls and age- and sex-matched controls without diarrhea. Using stool samples, gut microbiome was analyzed by whole-genome shotgun metagenomic sequencing, and gut metabolome was profiled by ultra-high performance liquid chromatography–mass spectrometry. Per-feature enrichment analyses of microbial species, metabolic pathways, and metabolites were done using multivariable linear models both cross-sectionally comparing MC to controls and longitudinally according to disease activity. Lastly, we performed multi-omics association analyses to assess the relationship between microbiome and metabolome data.

**Results:** We included 683 participants, 131 with active MC (66 with both active and remission samples), 159 with chronic diarrhea, and 393 age- and sex-matched controls without diarrhea. The stool microbiome in active MC was characterized by a lower alpha diversity as compared to controls and the remission phase of MC. Compared to controls, we identified eight enriched species in MC, most of which were pro-inflammatory oral-typical species, such as *Veillonella dispar* and *Haemophilus parainfluenzae*. In contrast, 11 species, including anti-inflammatory microbes such as *Blautia glucerasea* and *Bacteroides stercoris,* were depleted in MC. Similarly, pro-inflammatory metabolites, including lactosylceramides, ceramides, lysophospholipids, and lysoplasmalogens were enriched in active MC as compared to controls or MC cases in remission. Multi-omics association analyses revealed strong and concordant links between microbes, their metabolic pathways, and metabolomic profiles, supporting the tight interplay between disturbances in stool microbiome and metabolome in MC.

**Conclusion:** We observed a significant shift in stool microbial and metabolomic composition in MC. Our findings could be used in the future for development of non-invasive biomarkers for diagnosing and monitoring MC and developing novel therapeutics.

**What is already known on this topic:**

- Microbiome dysbiosis has been proposed to contribute to microscopic colitis (MC) pathogenesis.
- However, previous studies have been limited by small sample sizes, reliance on 16S rRNA sequencing technique, potential confounding by stool consistency, and lack of functional analyses of microbiome and longitudinal data. Moreover, the metabolomic composition of MC remain largely unknown.

**What this study adds:**

- In this largest longitudinal MC cohort with two control groups – chronic diarrhea controls and controls without diarrhea, gut microbiome of MC is characterized by a lower alpha diversity, enriched pro-inflammatory oral-typical species and depleted anti-inflammatory beneficial species.
- Gut metabolome of MC shows significant enrichment of pro-inflammatory metabolites, including lactosylceramides, ceramides, lysophospholipids, and lysoplasmalogens. Multi-omics analyses demonstrate strong and concordant relationships between microbes, metabolic pathways, and metabolomic profiles.

**How this study might affect research, practice or policy:**

- Our findings could facilitate development of non-invasive biomarkers and novel therapeutics for MC.

## Introduction

Microscopic colitis (MC) is an inflammatory bowel disease (IBD) characterized by chronic relapsing watery diarrhea. Although affiliated with the umbrella term of IBD, MC is distinct from ulcerative colitis (UC) / Crohn’s disease (CD) by lacking macroscopic inflammation that is visible under endoscopy. MC is one of the most frequent causes of chronic diarrhea, especially in women older than 65 years, and is associated with a significant impairment in quality of life [1]. Yet, the pathophysiology of MC is largely unknown. One proposed mechanism is that MC occurs as a result of inappropriate immune responses to environmental-induced perturbances in the gut microbiome [2]. This hypothesis is supported by the observation that fecal diversion can lead to endoscopic remission and reduction of pro-inflammatory cytokines, including TNF-α, while restoration of the gut continuity lead to disease recurrence and resurgence of pro-inflammatory cytokines [3]. Several smaller-scale studies have attempted to characterize the changes in the gut microbiome in MC and reported lower alpha diversity and a higher microbial dysbiosis index in patients with active MC compared to controls [4, 5]. Further cross-sectional taxonomic profiling have revealed reduced abundance of *Ruminococcaceae* [6], *Coriobacteriaceae* [7], *Clostridiales* [8], *Collinsella* [9], and *Akkermansia* [4, 10], and increased abundance of pro-inflammatory *Desulfovibrionales* [7] and *Prevotella* [8] comparing MC to controls.

Longitudinal data are scarce, with some reported decreased abundance of *Collinsella*, *Ruminococcaceae*, *Coriobacteriaceae* and *Clostridiales* in active MC compared to remission [6]. A recent study from our group using metagenomic sequencing of stool samples reported depletion of *Alistipes putredinis* in MC [5]. Despite these previous endeavors, research to-date has been limited by small sample sizes (MC participants: n = 10-52) [4, 5, 6, 7, 8, 9, 10], reliance on 16S rRNA sequencing techniques, which provides lower resolution data on composition and function of the gut microbiome [4, 6, 7, 8, 9], use of non-diarrhea controls only that could not account for confounding by stool consistency [4, 6, 7, 8, 9, 10, 11], lack of more direct data on function of the gut microbiome (e.g., metabolomics or transcriptomics) [4, 5, 6, 7, 9, 10] and limited data on differences in the gut microbiome according to disease activity [6, 7, 8, 9, 10].

Besides taxonomic composition and functional potential, the gut microbiome also contributes to and is influenced by the intestinal metabolomic landscape. In fact, some small molecules, including those derived from gut microbiome, such as short-chain fatty acids (SCFA), have been found to play an important role in host immune response [12]. However, few studies have analyzed the microbiome-metabolome correlations in patients with MC. Thus, we established a large longitudinal cohort study of patients with MC and two separate comparators – controls without diarrhea and individuals with chronic diarrhea to comprehensively characterize microbiome and metabolome alterations in MC, understand the longitudinal changes associated with disease activity, and identify potential microbiome or metabolome biomarkers for MC.

## Methods

### Study population

Starting January 2016, we invited all adults (≥ 18 years old) with chronic diarrhea who were seen for consultation or were scheduled for a diagnostic colonoscopy at the Massachusetts General Hospital (MGH) Gastroenterology unit to submit a stool sample and provide detailed information on their demographics and lifestyle factors (**Figure 1**). Inclusion criteria for the active MC group included: 1) presence of active symptoms (≥ 3 bowel movements/day and Bristol stool score [BSS] > 5) and 2) a confirmed histologic diagnosis of MC. The chronic diarrhea cohort consisted of individuals with active symptoms (≥ 3 bowel movements/day and BSS > 5) but a normal colonoscopy with biopsies. Patients with active MC were invited to provide a second stool sample during remission phase (< 3 bowel movements/day and BSS ≤ 5) at least 8 weeks after their initial stool sample submission. At baseline, we excluded individuals with diagnoses of UC or CD, colorectal cancer, prior colorectal surgery, and recent antibiotics use (within 3 months).

**Figure 1.**
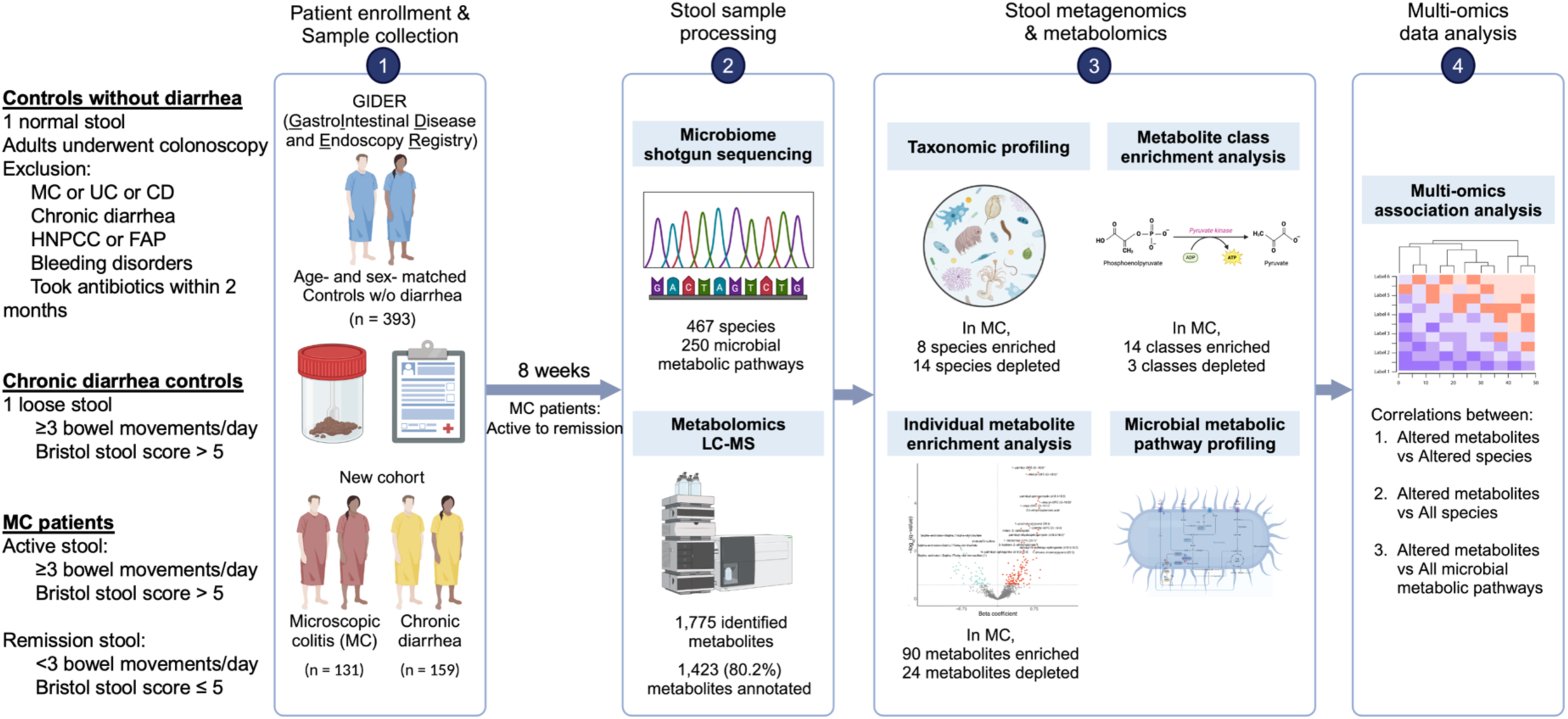
Overall study design. Illustration was produced with BioRender.com.

We also included individuals in our ongoing colonoscopy-based longitudinal cohort of GIDER (**G**astro**I**ntestinal **D**isease and **E**ndoscopy **R**egistry) [13], which included adult participants who were scheduled for a screening or surveillance colonoscopy at MGH (**Figure 1**). Before their colonoscopy, consented individuals were asked to complete a baseline medical history questionnaire and provide a stool sample. Participants with a history of UC or CD, hereditary non-polyposis colorectal cancer (HNPCC), familial adenomatous polyposis (FAP), recent antibiotics use (within 2 months), or known bleeding disorders were not eligible for this study. We used this cohort to identify non-diarrhea controls that were matched in a 3:1 ratio to active MC participants by age and sex. The study was approved by Partners Human Research Committee and the Institutional Review Board of Mass General Brigham (Protocol # 2015P001333 and # 2015P000275). All participants have provided informed consent before enrollment.

### Stool collection and processing

For all participants, consented individuals were provided with a stool specimen collection kit prior to their scheduled colonoscopy. Participants were instructed to collect the sample 1-2 days before the colonoscopy and either brought their pre-colonoscopy stool specimens at the time of their visit or shipped the specimens to our research facility. Stool samples were processed and frozen within 48 hours of collection. Participants provided stool specimens in storage tubes containing preservative RNAlater (Invitrogen, Fisher, Waltham, MA, USA) or 95% ethanol. Samples were handled at room temperature and stored at −80°C until nucleic acid extraction. Stool genomic DNA purification was performed using QIAGEN AllPrep MiniKit (Valencia, CA, USA) per the manufacturer’s instructions and consistent with the Human Microbiome Project protocols [14, 15].

### Metagenomic sequencing, taxonomic composition, and metabolic pathway profiling

Purified DNA from stool samples were processed using the Nextera XT DNA Library Preparation Kit (Illumina, San Diego, CA, USA) following manufacturer’s protocol to generate metagenomic sequencing libraries. Then, whole-genome shotgun metagenomic sequencing was performed at the Broad Institute (Cambridge, MA, USA) using Illumina NovaSeq 6000 Sequencing System with 150-base-pair paired-end reads. The raw metagenomic reads were processed using KneadData (v0.12.0) to remove Illumina adapters and short reads (< 50 base pairs). We used Bowtie2 (v2.5.1) to identify and remove reads that are of human origin based on the Genome Reference Consortium human genome template (GRCh37/hg19). After this, quality control of the metagenomes was conducted using FastQC (v0.12.0). During quality control, two samples with a read depth ≤ 7 million reads were removed. Four samples with unknown BSS were excluded, while seven chronic diarrhea samples were removed since initial collection was done after resolution of symptoms. After exclusion, 820 samples (out of 835 samples, 98.2%) were included in our study, with a post-quality control average read depth of 26.6 ± 9.8 million (∼ 3.7 Gbp per sample).

Taxonomic composition and functional pathways were profiled from metagenomes using the bioBakery meta’omics workflow [16]. The Metagenomic Phylogenetic Analysis tool (MetaPhlAn v4.0.6) was used to compare our samples’ shotgun sequences with the tool’s database to characterize taxonomies and species relative abundance in each sample [17]. We adopted the HMP Unified Metabolic Analysis Network (HUMAnN3 v3.8) to profile the relative abundance of functional pathways [18]. HUMAnN3 aligns the metagenomic sequences to two databases, ChocoPhlAn pangenome database [19] and UniRef90 protein database[20], to reconstruct the microbial metabolic pathway abundances based on species identified from MetaPhlAn. Finally, we limited our analysis to taxonomic species with relative abundance ≥ 0.01% and microbial metabolic pathways with relative abundance ≥ 0.1% that were present in ≥ 10% of samples [21].

### Metabolite profiling

Metabolomic measurements were conducted with ultra-high performance liquid chromatography–mass spectrometry (LC-MS) by Metabolon Inc. (Morrisville, NC, USA) in a subset of participants. Four LC-MS methods (Pos Late, Pos Early, Polar, and Neg) were performed. For analysis, we used the normalized imputed dataset from Metabolon, in which metabolites were rescaled by setting the median to 1, and missing values were imputed with the minimum value observed in each metabolite. Out of 1,775 identified metabolite features, 1,423 (80.2%) metabolites were annotated. For subsequent analyses, we limited metabolites to those annotated with relative abundance ≥ 0.1% and prevalence ≥ 10%.

### Metagenomic and metabolomic analysis

From the MetaPhlAn taxonomic outputs, we calculated the alpha diversity using Chao 1 index for each sample at species level. The average alpha diversity was then compared between the three groups using non-parametric Wilcoxon rank-sum tests. To compare the difference in microbiome composition across samples, we calculated the Bray-Curtis dissimilarity [22]. We performed classical multidimensional scaling with Bray-Curtis dissimilarity matrix and constructed principal coordinates analysis (PCoA) plots. Subsequently, permutational multivariate analysis of variance (PERMANOVA) was conducted with 999 permutations to quantify the proportion of observed variation in microbiome composition explained by patient factors including disease type, stool consistency as defined by BSS, and demographics (age, sex, and BMI). The analyses were performed using the R package Vegan (v2.6-4) [23].

In the cross-sectional analysis, to identify unique taxonomic features in MC, we compared relative abundance of species and pathways between MC versus non-MC controls using the Microbiome Multivariable Association with Linear Models (MaAsLin2 v3.18, http://huttenhower.sph.harvard.edu/maaslin2) [24], while adjusting for BSS (> 5 vs ≤ 5), age, sex, and BMI. Age and BMI were standardized into z-scores in the regression model by MaAsLin2. The linear model for cross-sectional analysis could be expressed as:

*Log (Microbiome (metabolite) features) ∼ intercept + Disease type (MC vs Chronic diarrhea vs Controls without diarrhea) + BSS (> 5 vs ≤ 5) + age + sex + BMI*

Metabolites were categorized into different classes based on Metabolon classification system, which assigns a metabolite class to each identified metabolite based on the relevant biochemical pathway. Using the same MaAsLin2 model, we compared the relative abundance of each metabolite in MC to controls. Metabolites altered in the same direction comparing MC to both controls were selected, and we then calculated and ranked the medians of t-statistics of metabolites within each metabolite class. Metabolite classes with a median > 0 were considered to be enriched in MC, while those with a median < 0 were considered depleted [25]. After identifying metabolite classes that were enriched and depleted in MC compared to both controls, we evaluated the abundance of all metabolites in the class cross-sectionally.

For the longitudinal comparison, we limited our analyses to species and metabolites that were significantly different between MC and both controls and used Wilcoxon signed-rank tests to compare the relative abundances between active and remission samples. In exploratory analysis, we evaluated whether our observed differences in relative abundances of species and metabolites were consistent across the histologic subtypes (lymphocytic colitis (LC) versus collagenous colitis (CC)) of MC. For all analyses, multiple testing corrections were implemented using the Benjamini-Hochberg method when appropriate, and a false discovery rate (FDR) q < 0.25 was considered statistically significant, consistent with prior discovery-based analysis of microbiome and metabolomics data [21]. All p-values were two-sided.

### Microbe-metabolite-microbial metabolic pathway correlation analysis

To understand the extent of metabolome alterations attributable to dysbiosis in MC, we performed hierarchical correlation analyses between microbiome compositions, microbial metabolic pathways, and metabolomic profiles using HAllA (Hierarchical All-against-All association testing) [26]. HAllA reports significant Spearman’s correlation coefficient (rho) between paired large multi-omics datasets, with statistical significance determined by permutations. Two correlation analyses were conducted. First, to determine if our observed metabolite alterations were linked to perturbations in species composition, we calculated the correlations of altered metabolites against all microbes and reported metabolite-microbe correlations. Second, to evaluate whether the metabolite alterations may be attributable to microbial metabolism, we estimated the correlations between altered metabolites and microbial metabolic pathways and reported metabolite-pathway correlations. For all the correlation analyses, we only reported those with Spearman’s rho > 0.15 or < −0.15, consistent with prior metabolite-microbe correlation analysis [27].

## Results

### Patients with MC have a pro-inflammatory and aerotolerant microbiome

After quality control, we enrolled 683 participants, including 131 patients with active MC, 159 patients with chronic diarrhea, and 393 age- and sex-matched controls without diarrhea for metagenome analysis (**Table 1**). Individuals with MC were in average older, more likely to be female, and had lower BMI as compared to chronic diarrhea controls. Our longitudinal cohort included 66 pairs of stool samples from patients with MC in both active and remission phase, among which 27 had CC and 33 had LC.

**Table 1.** Baseline characteristics of participants with microscopic colitis (MC) and comparator groups.

|  | **Active MC**<br>**N = 131** | **Chronic Diarrhea**<br>**N = 159** | **Controls w/o diarrhea**<br>**N = 393** |
| --- | --- | --- | --- |
| Age (years), mean (SD) | 62.9 (14) | 55.5 (16) | 62.5 (11) |
| Female (%) | 109 (83) | 111 (70) | 327 (83) |
| BMI (kg/m<sup>2</sup>), mean (SD) | 24.9 (4.7) | 28.6 (7.0) | 26.2 (5.5) |
| Race, n (%) |  |  |  |
| White | 123 (98) | 148 (95) | 364 (92) |
| Other | 2 (2) | 7 (5) | 29 (8) |
| Smoking, n (%) |  |  |  |
| Current | 9(8) | 3 (4) | 13 (4) |
| Never | 56(53) | 47 (64) | 217 (58) |
| Past | 41(39) | 24 (32) | 143 (38) |
| Antibiotics use in the past 12 months, n (%) | 39 (31) | 49 (34) | 94 (25) |

For the overall composition of the gut microbiome, we observed that the alpha diversity (Chao 1 index) was significantly lower in active MC as compared to controls without diarrhea (Wilcoxon p < 0.001) but not to chronic diarrhea controls (Wilcoxon p = 0.446) (**Figure 2A**). Similarly, in longitudinal analysis, the gut microbiome in active MC had a lower alpha diversity as compared to remission (Wilcoxon p = 0.033) (**Figure 2B**). Similar trends were observed in subgroup analysis for subtypes of MC (**Fig S1A-C**). Principal coordinate analysis (PCoA) based on the Bray-Curtis dissimilarity of microbial composition showed no significant difference between the structure of the gut microbiome according to disease status or activity (**Figure 2C-D**). These findings were corroborated in our PERMANOVA analysis where both disease type (MC vs chronic diarrhea vs control without diarrhea) and disease activity (active vs remission) explained 2.2% and 1.8% of the variation observed in stool microbiome composition, but neither reached statistical significance (**Fig S2**).

**Figure 2.**
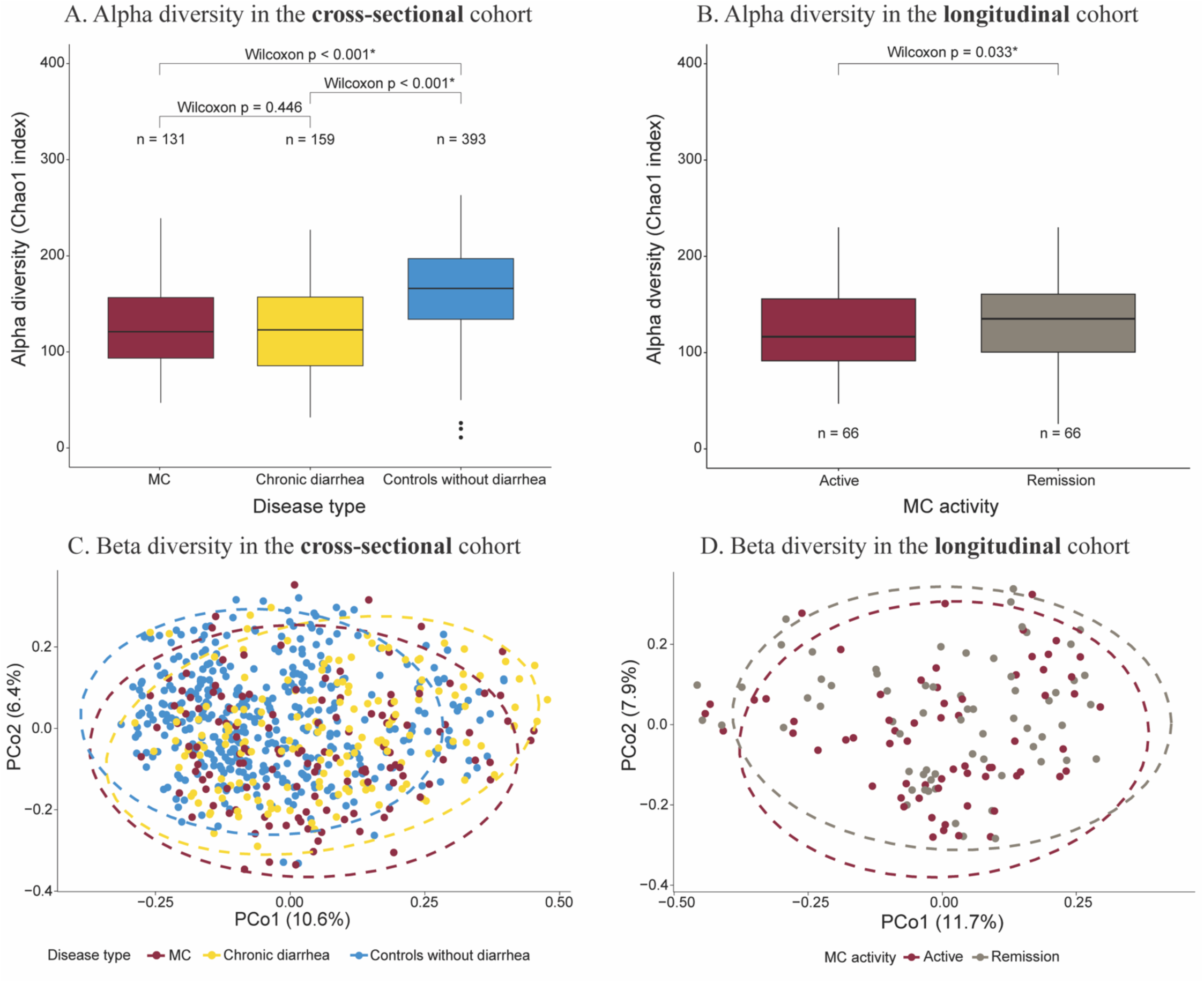
Differences between microbial composition of MC and comparator groups. (A, B) Alpha diversity (Chao 1 index) in the cross-sectional (A) and longitudinal cohorts (B). Cross-sectionally, the alpha diversity in the MC microbiome was lower than that of controls without diarrhea and similar to the chronic diarrhea group. Longitudinally, the alpha diversity was higher in the remission phase compared to active phase among patients with MC. (C, D) Principal coordinate analysis (PCoA) based on Bray-Curtis dissimilarity showed no significant differences in microbial composition comparing MC versus chronic diarrhea or controls without diarrhea (C) and active MC versus MC in remission (D).

In feature-level analyses for the relative abundance of taxonomic species, we included 467 microbial species after quality control and filtering the metagenomic reads (relative abundance ≥ 0.01% and prevalence ≥ 10%). In our cross-sectional analysis, compared to controls without diarrhea, patients with MC had 46 enriched species and 234 depleted species (Benjamini– Hochberg FDR q < 0.25) while compared to chronic diarrhea, patients with MC had 36 enriched species and 17 depleted species (Benjamini–Hochberg FDR q < 0.25, **Fig S3**). Eight species were enriched in MC compared to both controls, while 11 species were depleted (**Figure 3A**). Among the enriched species, *Veillonella dispar*, *V. parvula*, *V. rogosae*, and *Haemophilus parainfluenzae* are pathologic oral bacteria which have also been shown to be enriched in IBD [28, 29, 30]. Similarly, several depleted species, including *Collinsella spp.*, *Mediterraneibacter butyricigenes*, *Firmicutes spp.*, *Blautia glucerasea*, and *Bacteroides stercoris* have also been showed to be decreased in IBD [31, 32, 33, 34, 35]. Notably, many of these species are known to have anti-inflammatory properties (e.g., *B. stercoris* [35]) or produce metabolites such as SCFA (e.g. *B. glucerasea* [34]) that can suppress inflammation. We then compared the top differentially abundant species identified cross-sectionally in the longitudinal cohort, and the results showed seven species were depleted in active MC compared to MC in remission, while no species were enriched (**Figure 3C**). Six out of the seven depleted species (except *Nitrosopumilus_SGB14899*) in active disease were obligate anaerobes (such as *Clostridiales_bacterium_NSJ_32* and *Collinsella_SGB4121*), supporting the notion that in the pro-inflammatory state there is excessive oxidative stress [30, 36]. In exploratory analysis, we showed that most MC-enriched species were consistently enriched in both LC and CC compared to controls while MC-depleted species were only consistently depleted in CC (**Fig S4**). This finding suggests that there may be unique differences in the gut microbiome composition according to histologic subtypes of CC and LC.

**Figure 3.**
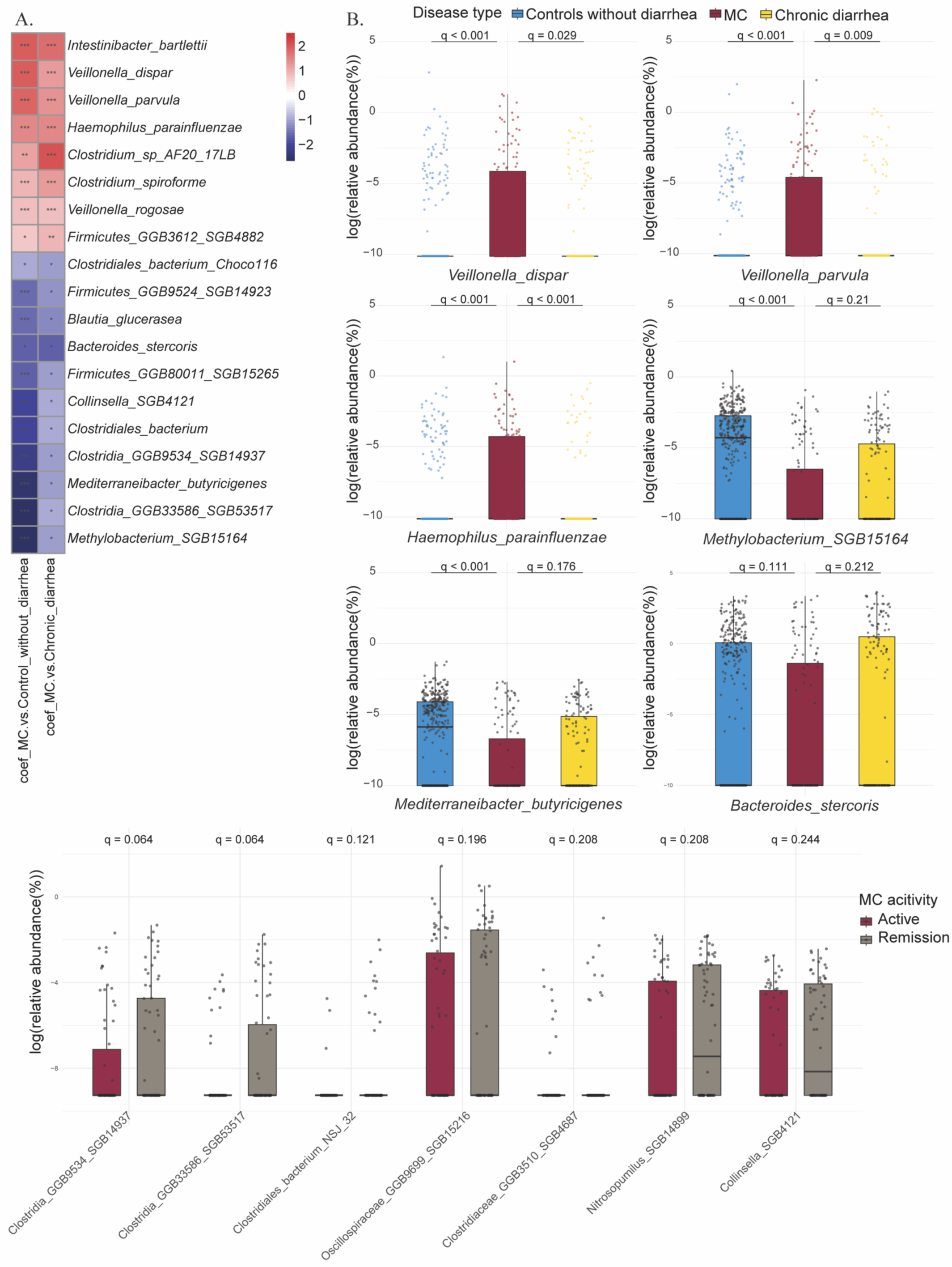
Comparisons of relative abundance of altered species according to disease type and MC activity. (A) The heatmap shows the β coefficient of species with altered abundance comparing MC to both controls in the cross-sectional cohort. We used the following model to adjust for confounding factors: *Log (Microbiome features) ∼ intercept + <u>Disease type (MC vs Chronic diarrhea vs Controls without diarrhea)</u> + BSS (> 5 vs ≤ 5) + age + sex + BMI*. *** FDR q < 0.05; ** 0.05 ≤ FDR q < 0.1; * 0.1 ≤ FDR p < 0.25. (B) Examples of log-transformed abundances of altered species in MC group in the cross-sectional cohort. *V. dispar*, *V. parvula*, and *H. parainfluenzae* were enriched in MC, while *Methylobacterium_SGB15164*, *M. butyricigenes*, and *B. stercoris* were depleted. Data were shown as median with interquartile range (IQR, 25^th^ and 75^th^). Whiskers indicated 1.5*IQR. (C) Examples of log-transformed abundances of altered species in active MC versus MC in remission in the longitudinal cohort. All species were depleted in active MC compared to remission. Benjamini-Hochberg FDR q-values were derived from Wilcoxon signed rank tests.

### Patients with MC have a pro-inflammatory metabolome and disturbed lipid metabolism

We profiled the gut metabolome in 335 patients, including 92 patients with active MC, 138 patients with active chronic diarrhea, and 105 controls without diarrhea. In the longitudinal cohort, all 33 MC patients had metabolome data. The PCoA plot for metabolite composition revealed more distinct distributions of MC vs control without diarrhea group compared to the PCoA plot of microbial composition (**Fig S5A**). Moreover, we found strong positive correlation (Spearman rho = 0.579, p < 0.001) between the first axes of ordination of metabolome and microbiome datasets, suggesting strong coupling of microbial composition, metabolic profile, and disease type. Similarly in the longitudinal cohort, we observed greater distinction between active versus remission phase of MC by metabolite composition than microbiome (**Fig S5B**). Specifically, there was a strong negative correlation (Spearman rho = −0.550, p < 0.001) between metabolome and microbiome, suggesting that while there is a decrease in alpha diversity in the active phase of MC, the metabolic activity is significantly increased during this phase.

Enrichment analyses were performed for the 1,423 known metabolites using MaAsLin2. We first compared the abundance of metabolite classes in MC versus each control group (**Fig S6A-B**). Among the altered metabolite classes, 14 were enriched and three were depleted in MC as compared to both control groups (**Figure 4A**). Top enriched metabolite classes included lactosylceramides (LCER), long chain polyunsaturated fatty acids (LC-PUFAs), lysoplasmalogens, fatty acid metabolism (acyl carnitine, long chain saturated) metabolites, ceramides, and lysophospholipids. Top depleted metabolite classes included fatty acid/dicarboxylate, sterol, and androgenic steroids. Many of these metabolite classes have also been shown to be altered in IBD [12]. Within the altered metabolite classes, there were 114 altered individual metabolites (**Figure 5A and Table S1**). Among the top metabolite classes, we conducted detailed enrichment analyses of individual metabolites for the four classes – LCER, ceramides, lysophospholipids, and lysoplasmalogens based on their consistent results when comparing to both controls and prior literature suggesting potential biological role in other IBD [12, 37, 38] and immune-mediated diseases [39]. In our detailed enrichment analyses, the total levels of LCER (n = 3), ceramide (n= 5), lysophospholipid (n = 14), and lysoplasmalogen (n = 8) metabolites were elevated in active MC compared to the two control groups and the remission phase of MC (**Figure 4B-E**). In addition, we also found elevated levels of individual metabolites from each of the four metabolite classes in active MC, including lactosyl-N-palmitoyl-sphingosine (d18:1/16:0), N-palmitoyl-sphingosine (d18:1/16:0), 1-stearoyl-GPC (18:0), and 1-stearyl-GPE (O-18:0), which have all been showed to be enriched in IBD and other immune-mediated diseases [12, 37, 38, 39] (**Figure 5B-E**). Overall, the metabolomics data revealed profound disturbance of lipids (sphingolipids and phospholipids) metabolism in active MC.

**Figure 4.**
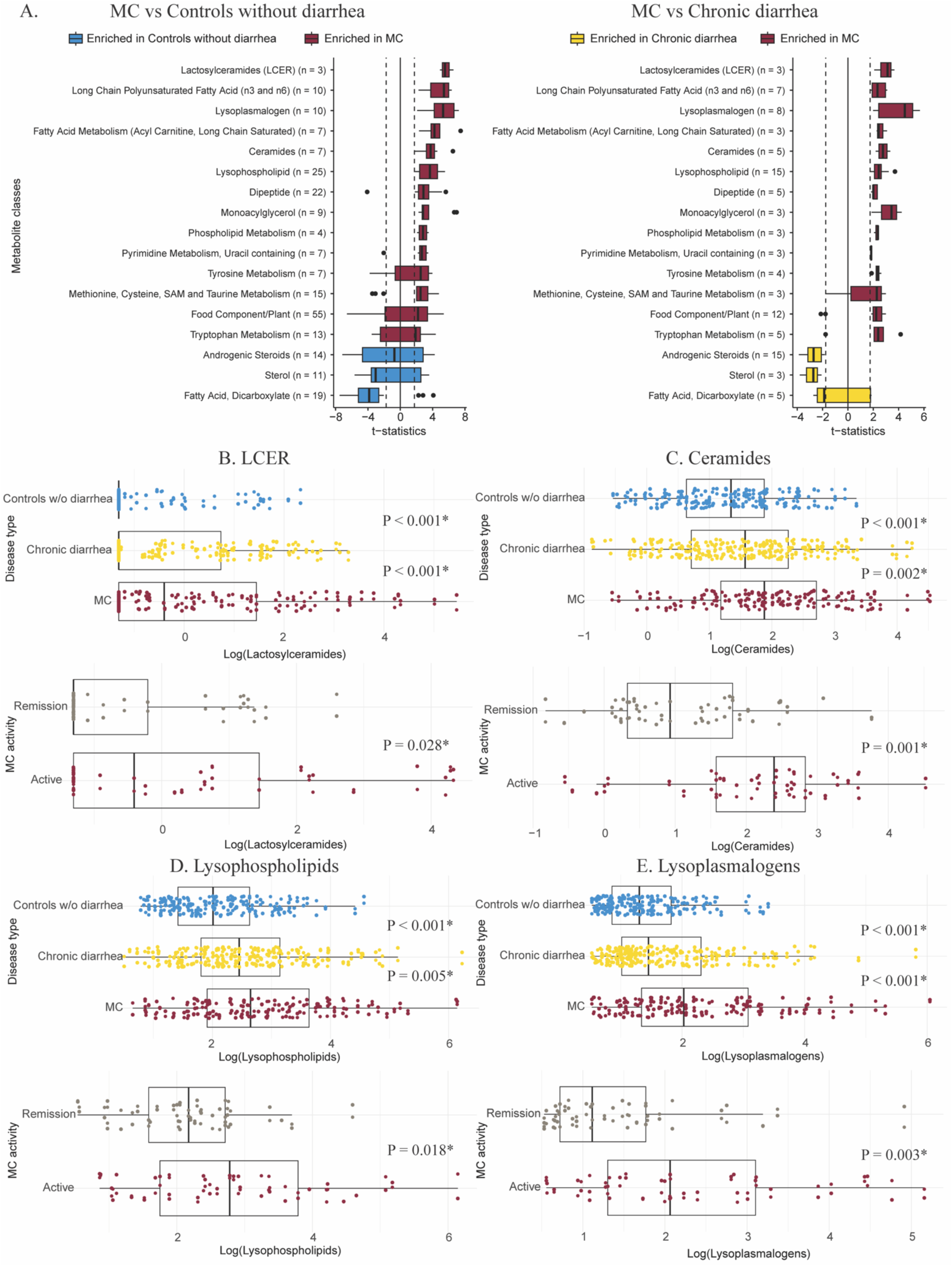
Comparisons of relative abundance of altered metabolite classes according to disease type and MC activity. (A) The box plots showed that 14 metabolite classes were enriched (median t-statistics > 0) and three metabolite classes were depleted (median t-statistics < 0) in MC compared to both controls after multi-variable adjustment. The dotted lines illustrated the significance threshold of an individual metabolite. T-statistics were derived from MaAsLin using the model: *Log (Metabolite features) ∼ intercept + <u>Disease type (MC vs Chronic diarrhea vs</u> <u>Controls without diarrhea)</u> + BSS (> 5 vs ≤ 5) + age + sex + BMI*. (B-E) Enrichment analyses of metabolite class in the cross-sectional (top) and longitudinal cohorts (bottom). We used the following model to adjust for confounding factors in the longitudinal cohort: *Log (Metabolite features) ∼ intercept + <u>BSS (> 5 vs ≤ 5)</u> + age + sex + BMI*.

**Figure 5.**
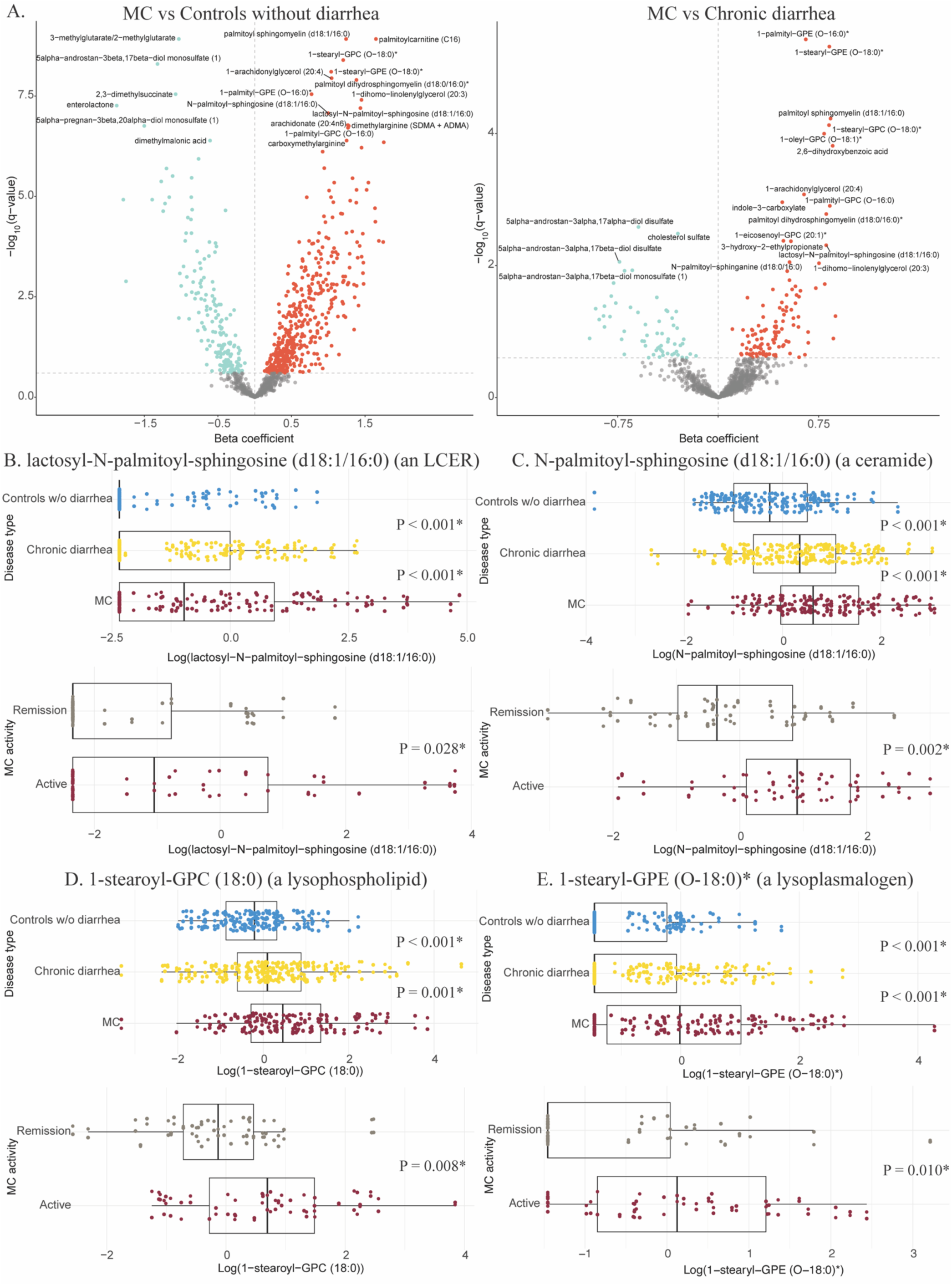
Comparisons of relative abundance of altered individual metabolites according to disease type and MC activity. (A) The volcano plots demonstrated alteration of individual metabolites’ relative abundance in MC compared to both controls. Red color indicated enriched relative abundance, turquoise color indicated depleted relative abundance, and gray color indicated no change in relative abundance. Coefficients and q-values were derived from MaAsLin using the same model. (B-E) Enrichment analyses of individual metabolite from each of the four metabolite classes in the cross-sectional (top) and longitudinal cohorts (bottom).

LCER and ceramides are components of the broader sphingolipid metabolism pathway (**Fig S7**), known to be enriched in IBD [25]. Besides LCER and ceramides, we identified another four MC-enriched metabolites in this pathway among the 114 altered metabolites, including palmitoyl sphingomyelin (d18:1/16:0) (a sphingomyelin), N-palmitoyl-sphinganine (d18:0/16:0) (a dihydroceramide), palmitoyl dihydrosphingomyelin (d18:0/16:0) (a dihydrosphingomyelin), and glycosyl-N-palmitoyl-sphingosine (d18:1/16:0) (a glucosylceramide) (**Figure 4B**). In exploratory analysis, we examined the differences in the metabolic profiles between MC and controls according to histologic subtypes of CC and LC and observed similar patterns (**Fig S8**).

### Links between microbiome composition, microbial metabolic pathways, and metabolomic profiles

To understand how gut microbiome may alter the metabolomic landscape, we tested the correlations between altered species and altered metabolites (**Figure 6A**). Overall, we saw a strong correlation between perturbed microbes and altered metabolites in MC, suggesting that our observed metabolic changes is likely linked to compositional changes in the gut microbiome. We used sphingolipids (**Figure 6B**) to illustrate the strong concordance between species and metabolites. Levels of sphingolipid metabolites, which were elevated in MC, were positively correlated with relative abundance of *V. parvula*, which was enriched in MC (**Figure 6C**). Conversely, levels of sphingolipids metabolites were negatively correlated with relative abundance of *Methylobacterium_SGB15164*, which was depleted in MC (**Figure 6D**). We also considered the possibility that metabolomic disturbance in MC may be related to dynamic changes in the function of the gut microbiome and therefore additionally evaluated the correlations between: (1) all microbial species and altered metabolites and (2) all microbial metabolic pathways and altered metabolites.

**Figure 6.**
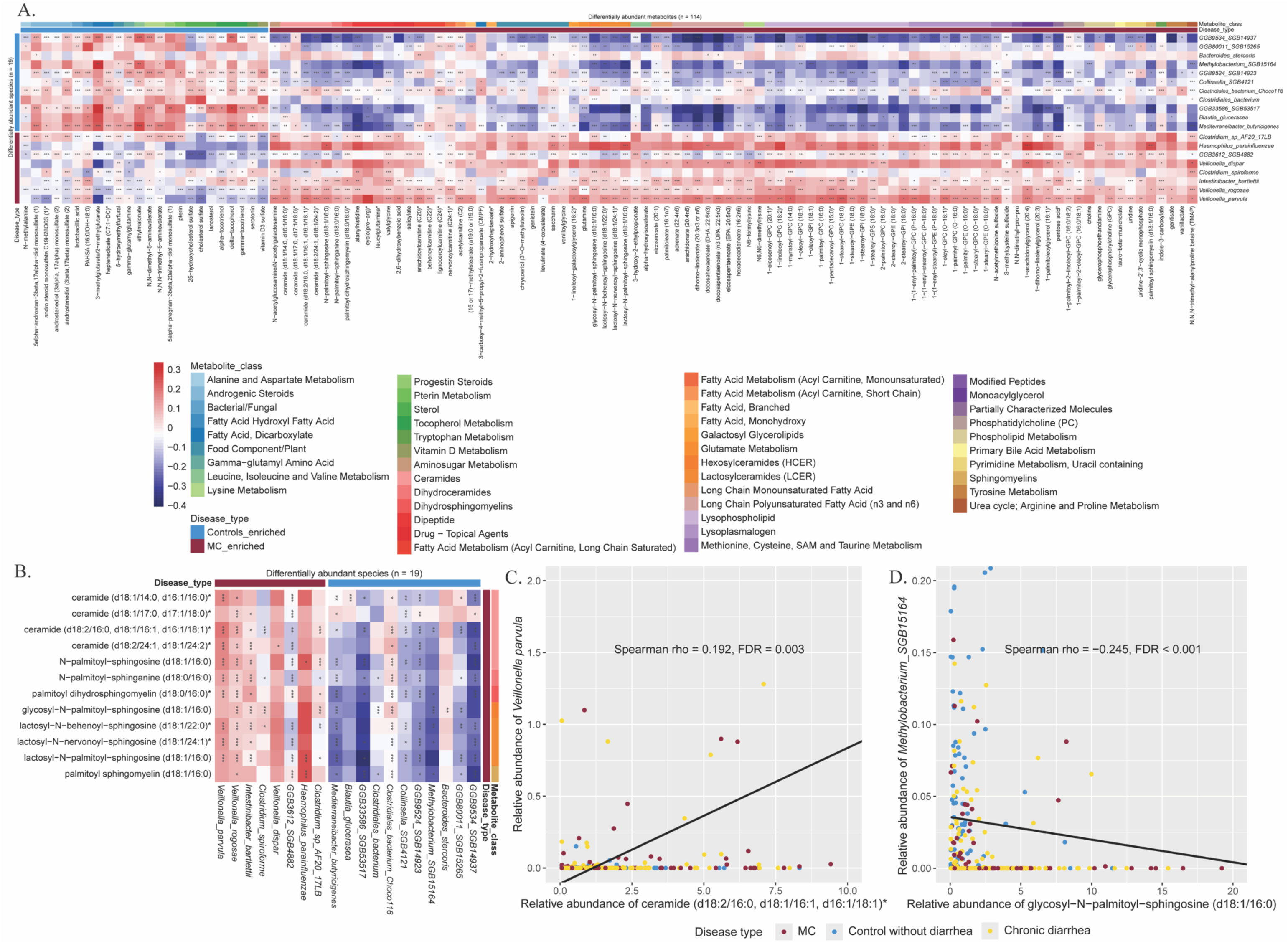
Correlations between differentially abundant species and metabolites grouped by disease type (MC vs both controls). (A) Metabolites and species were classified and clustered as MC-enriched (brown) or control-enriched (blue, including controls without diarrhea and chronic diarrhea), and metabolites were further grouped by metabolite class. The abundance of metabolites and microbes covariate concordantly. There was a clear pattern that MC-enriched species and metabolites (or control-enriched species and metabolites) were positively correlated, while MC-enriched species and control-enriched metabolites (or control-enriched species and MC-enriched metabolites) were negatively correlated. *** FDR q < 0.05; ** 0.05 ≤ FDR q < 0.1; * 0.1 ≤ FDR p < 0.25. (B) Correlations between sphingolipids (MC-enriched metabolite) and species. (C, D) Examples of individual correlations between MC-enriched sphingolipids and *V. parvula* (MC-enriched) or *Methylobacterium SGB15164* (control-enriched). Concordantly, positive correlations were observed between MC-enriched species and MC-enriched sphingolipids, while negative correlations were found between control-enriched species and MC-enriched sphingolipids.

For correlations between all microbial species and altered metabolites, we reported the species highly correlated with sphingolipids, lysophospholipids, and lysoplasmalogens, all of which are pro-inflammatory (**Fig S9A-C**). From these correlations, we highlighted the key species, which have altered abundance in MC in our study or have been reported to be associated with MC in literature, and produced simplified heatmaps (**Fig S10A-C**). Concordantly, among species with altered abundance (**Figure 3A**), pathologic species (such as *H. parainfluenzae* and *Veillonella spp*.) were more abundant in MC and in gut with elevated pro-inflammatory metabolites, while protective species (such as *Methylobacterium_SGB15164*, *M. butyricigenes*, and *B. glucerasea*) were depleted in MC and in increased pro-inflammatory metabolites (**Fig S11A-C**). The concordance between differential abundance analysis and functional analysis suggested that pathologic species may produce pro-inflammatory metabolites in MC, or pro-inflammatory metabolites produced by MC gut may inhibit the growth of protective species. Although not observed in our differential abundance analysis, a number of notable species that have previously been linked to MC or IBD were also functionally correlated with the altered metabolites in our study. For instance, *Ruminococcus gnavus*, which has been reported to be strongly enriched in IBD [40], was positively correlated with sphingolipids, lysophospholipids, and lysoplasmalogens, while *Akkermansia spp.,* which have been shown to be depleted in IBD and MC [4, 10], were negatively correlated with these metabolites.

For correlations between all microbial metabolic pathways and altered metabolites, we presented the pathways highly correlated with sphingolipids, lysophospholipids, and lysoplasmalogens (**Fig S12A-C**). Several important pathways were correlated with elevated levels of pro-inflammatory metabolites. Enriched sphingolipids were correlated with altered carbohydrate metabolism (increased degradation and decreased synthesis of sugar) in bacteria, increased pentose phosphate pathway and purine/pyrimidine synthesis and salvage (DNA/RNA synthesis), upregulated peptidoglycan maturation (bacterial cell wall) and phospholipid synthesis (bacterial cell membrane), suggesting some bacteria may proliferate faster in inflamed gut (**Fig S13A**). Lysophospholipid levels were positively correlated with phospholipid synthesis pathway (the direct upstream metabolite of lysophospholipids) in bacteria, implicating that some pathologic species may produce pro-inflammatory lysophospholipids that may be linked to inflammation in MC (**Fig S13B**). Elevated levels of lysoplasmalogens were correlated with downregulated glycogen synthesis (**Fig S13C**), likely suggesting increased environmental stress during active MC phase [41].

## Discussion

In this large cohort study, we comprehensively characterized the composition of the stool microbiome and metabolome in patients with MC compared to chronic diarrhea and controls without diarrhea. Consistent with prior work from our group and others [4, 5], we broadly saw a decrease in microbiome richness in active MC as compared to controls without diarrhea and the remission phase of the disease. In per-feature analysis, we found enrichment in pathogenic oral-typical species such as *H. parainfluenzae* and *Veillonella spp*. in the gut of MC patients as compared to controls, while species with anti-inflammatory properties such as *B. glucerasea* and *B. stercoris* were reduced in relative abundance in MC. In fact, oral-typical species have been proposed to be capable of translocating to the intestine and directly exacerbate IBD [42]. Our analysis revealed a marked increase in pro-inflammatory metabolites such as LCERs, ceramides, lysophospholipids, and lysoplasmalogens. Lastly, multi-omics analysis revealed a strong functional association across the microbiome and metabolome suggesting that compositional changes in the gut microbiome closely correlate with structural alterations in gut metabolome in MC.

Our findings are in line with prior studies of the gut microbiome in IBD. Specifically, consistent with our findings, pro-inflammatory species such as *Intestinibacter bartlettii*, *Veillonella spp.*, and *H. parainfluenzae* have been widely reported to be enriched in patients with IBD [28, 29, 30, 42, 43, 44], while species such as *C. bacterium*, *M. butyricigenes*, *B. glucerasea*, *B. stercoris* have been shown to be depleted in the gut microbiome of patients with IBD or MC [8, 9, 32, 35]. Interestingly and in contract to prior studies in IBD, we did not observe an association with *Ruminococcaceae* [6], *Coriobacteriaceae* [7], *Akkermansia* [4, 10], *Desulfovibrionales* [7], *Prevotella* [8], or *Alistipes* [5]. This may in large be explained by our adoption of multiple control groups, eliminating confounding effects by stool consistency, which has previously been shown may lead to false association with species such as *Ruminococcaceae*, *Akkermansia*, and *Prevotella* [11]. No prior study has examined stool metabolites in MC; however, our results were highly consistent with those previously reported in IBD [12, 45, 46].

Our study extends the current knowledge built on prior research. First, through comparison to multiple control groups and longitudinal sampling according to disease activity, we were able to identify gut microbiome compositional and functional changes that are likely unique to MC. Second, we identified a number of key metabolites that are disturbed in MC. For instance, lactosyl-N-palmitoyl-sphingosine (d18:1/16:0) (an LCER), N-palmitoyl-sphingosine (d18:1/16:0) (a ceramide), 1-stearoyl-GPC (18:0) (a lysophospholipid), and 1-stearyl-GPE (O-18:0)* (a lysoplasmalogen) are highly enriched in active MC relative to controls and those in remission. Third, through correlation analysis, we demonstrated that many of perturbations in the gut microbiome composition are closely linked to gut metabolome, suggesting that the shift in the gut microbiome likely have biological significance. Lastly, we explored differences in the gut microbiome and metabolome according to histologic subtypes of MC. Although the gut metabolomic landscapes were similar across CC and LC, we found that more species in the CC microbiome were significantly disturbed than those in LC microbiome, indicating the gut microbiome compositions may be distinct between MC subtypes.

Our findings are biologically plausible. In MC, an inflammatory milieu is expected to increase oxidative stress which in turn may lead to reduction in obligate anaerobic bacteria such as *Collinsella spp*. and *Clostridiales bacterium* with anti-inflammatory properties. Similarly, ectopic transplantation of oral microbes in the intestine has been shown to induce T helper 1 (T_H_1) cells and gut inflammation ^30^. In our metabolomics analysis, sphingolipids were highly enriched in active MC, most of which were human-derived and pro-inflammatory [25, 47]. For example, LCERs, also known as CD17 on leukocytes, promote inflammation through releasing arachidonic acids, facilitating neutrophil adhesion, and generating reactive oxygen species [48, 49]. Similarly, ceramides can induce inflammation by activating NF-κB and COX-2 pathway [50] or by being degraded to sphingosine and sphingosine-1-phosphate (S1P), which can modulate lymphocyte migration from lymph nodes [12, 51]. Lastly, lysophospholipids, which are enriched in active MC, can increase intestinal epithelial barrier permeability, facilitate bacterial translocation, and promote pathological Th1-mediated intestinal inflammation [38, 52, 53].

Our metabolomics findings have the potential to identify novel biomarkers for diagnosing and monitoring MC [12, 54]. Additionally, the fact that ceramides were substantially enriched in active MC indicated that S1P receptor modulation by for S1P inhibitors could potentially be a promising therapeutic target [55, 56, 57]. Although we did not directly identify S1P in our metabolomics profiling, its upstream metabolite sphingosine and downstream metabolite palmitate were successfully found and quantified. Patients with MC had elevated levels of sphingosine compared to controls without diarrhea, but the levels were similar to those of chronic diarrhea. In the longitudinal cohort, sphingosine levels were significantly higher in active MC compared to remission (**Fig S14A**). Similar trends were also found in palmitate analysis (**Fig S14B**).

Our study has several strengths. First, our largest sample size to date with longitudinal follow-up enabled us to compare the microbial and metabolic compositions according to disease type and activity and explore the difference between LC and CC. Second, by adopting multiple control groups, we decreased the possibility of confounding by stool consistency [11]. Third, the whole-genome shotgun metagenomic sequencing method provided higher resolution data on microbiome, generating species-level taxonomic profiles as compared to 16S rRNA sequencing, which generally only provides genus-level data [4, 6, 7, 8, 9], but more importantly, cannot be mapped against curated databases for metabolic pathway reconstruction [58]. Forth, we simultaneously analyzed the microbiome and metabolome, allowing us to examine both the abundance and functional changes in the gut microbiome to cross-validate multi-omic signatures of inflammation. Moreover, through additional analyses, we were able to assess the functional significance of microbiome changes as it relates to metabolic activity. We acknowledge several limitations. First, we did not systematically collect diet, medication, and lifestyle information, which could have confounded our observed associations. Second, our participants were mostly White, which could limit the generalizability of our findings to other populations. Lastly, we acknowledge that the observational nature of our study may limit our ability to make any conclusion about the biological roles of the observed changes in the gut microbiome and metabolome in the pathophysiology of MC.

In conclusion, patients with MC have dysbiotic gut microbiome and pro-inflammatory metabolome. Subtypes of MC, LC and CC, were subtly distinct in microbial and metabolic compositions. In addition to deepening our understanding of MC pathogenesis, our findings have potential clinical implications including helping to identify non-invasive biomarkers for diagnosing and monitoring MC and offering potential clues for novel therapeutic targets for MC.

## Data Availability

All data produced in the present study are available upon reasonable request to the authors.

## Abbreviations

BMI: Body mass index
CC: Collagenous colitis
CD: Crohn’s disease
FAP: Familial adenomatous polyposis
FDR: False discovery rate
GIDER: GastroIntestinal Disease and Endoscopy Registry
HAllA: Hierarchical All-against-All association testing
HNPCC: Hereditary non-polyposis colorectal cancer
IBD: Inflammatory bowel disease
ICAM-1: Intercellular cell adhesion molecule-1
LC: Lymphocytic colitis
LCER: Lactosylceramides
LC-MS: Liquid chromatography–mass spectrometry
LC-PUFAs: Long chain polyunsaturated fatty acids
LPC: Lysophosphatidylcholine
LPE: Lysophosphatidylethanolamine
LPS: Lysophosphatidylserine
MC: Microscopic colitis
MaAsLin2: Microbiome Multivariable Associations with Linear Models
NF-κB: Nuclear factor kappa B
PCoA: Principal coordinates analysis
PERMANOVA: Permutational multivariate analysis of variance
S1P: Sphingosine-1-phosphate
SCFA: short-chain fatty acids
TNF-α: Tumor necrosis factor alpha
UC: Ulcerative colitis

## Author contributions

Study design and concept: HKh

Study funding: DCC, HKh and RJX

Acquisition of data: DCC, HKh, JG, JM, KW, and RJX

Statistical analysis: ASYC, EN, HKi, JS, LN

Drafting of the manuscript: ASYC and HKh

Critical revision of the manuscript: All authors

## Data Sharing

The data that support the findings of this study are available upon reasonable request.

## Ethical Approval

The study was approved by Partners Human Research Committee and the Institutional Review Board of Mass General Brigham (Protocol # 2015P001333 and # 2015P000275).

## Transparency declaration

The lead author (the manuscript’s guarantor) affirms that the manuscript is an honest, accurate, and transparent account of the study being reported; that no important aspects of the study have been omitted; and that any discrepancies from the study as originally planned have been explained.

