## Supplementary material for "Composition and Function of the Gut Microbiome in Microscopic Colitis": Fig S1

**S figure 1.** Alpha diversity (Chao 1 index) in stool samples from different groups.

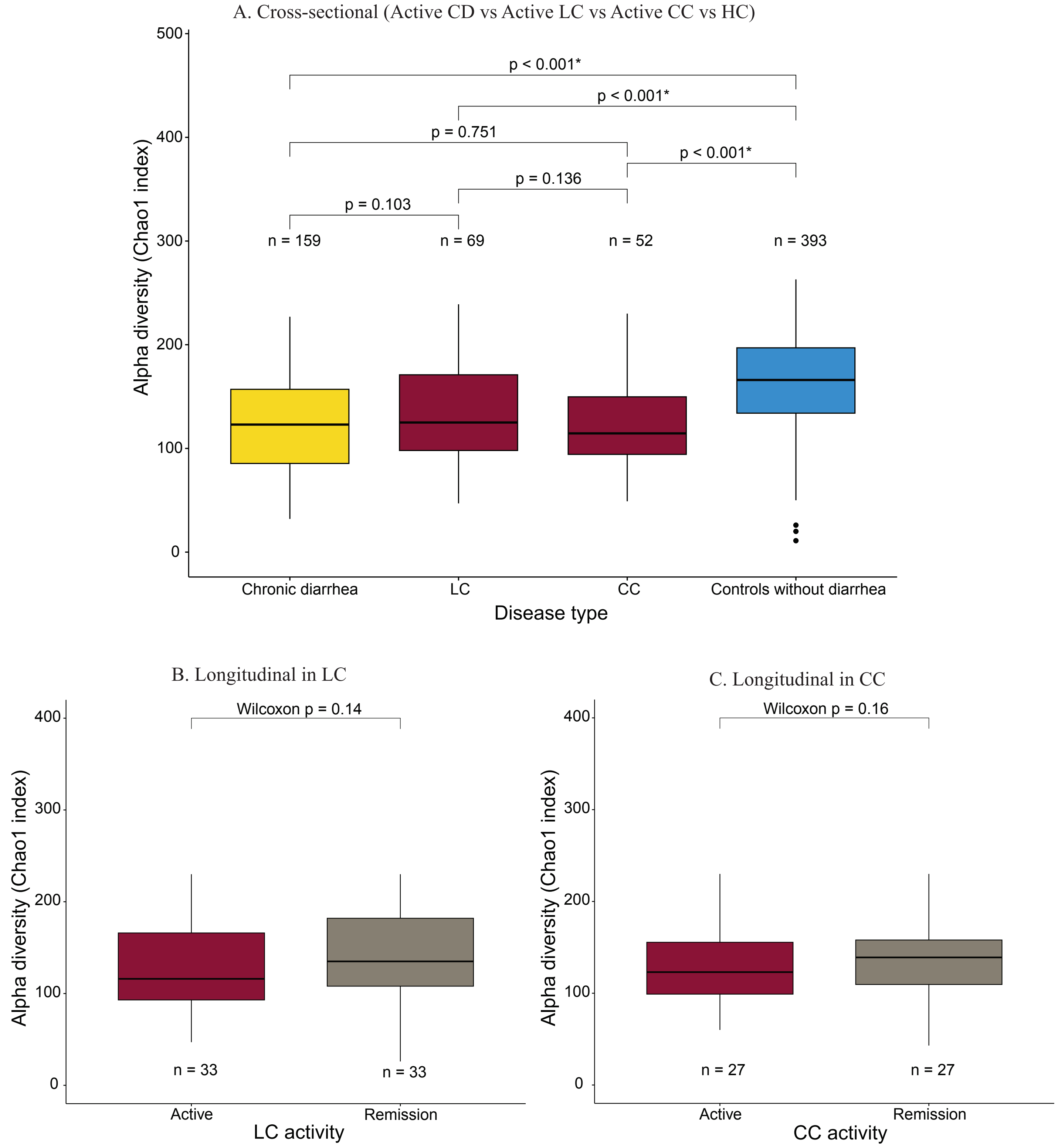


(A) Alpha diversity (Chao 1 index) in the cross-sectional cohort, with MC divided into LC and CC. The alpha diversities in the LC and CC microbiome were lower than that of controls without diarrhea and similar to the chronic diarrhea group.

(B, C) Longitudinally, the alpha diversity was higher in the remission phase compared to active phase among patients with LC (B) and CC (C). However, neither reached statistical significance.
