## Supplementary material for "Composition and Function of the Gut Microbiome in Microscopic Colitis": Fig S2

**S figure 2.** Proportion of variation in microbiome composition explained by individual factors based on PERMANOVA of the Bray-Curtis dissimilarity matrix.

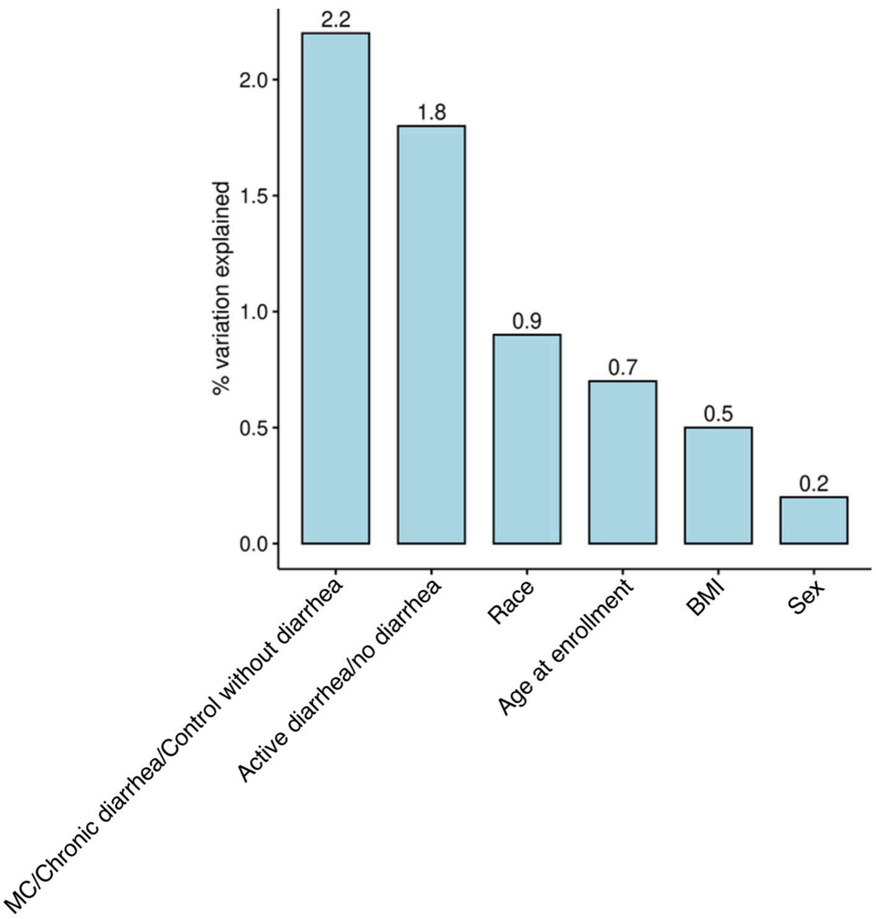
