## Supplementary material for "Composition and Function of the Gut Microbiome in Microscopic Colitis": Fig S3

**S figure 3.** Differential abundance analysis of species in MC compared to HC and CD in the cross-sectional cohort.

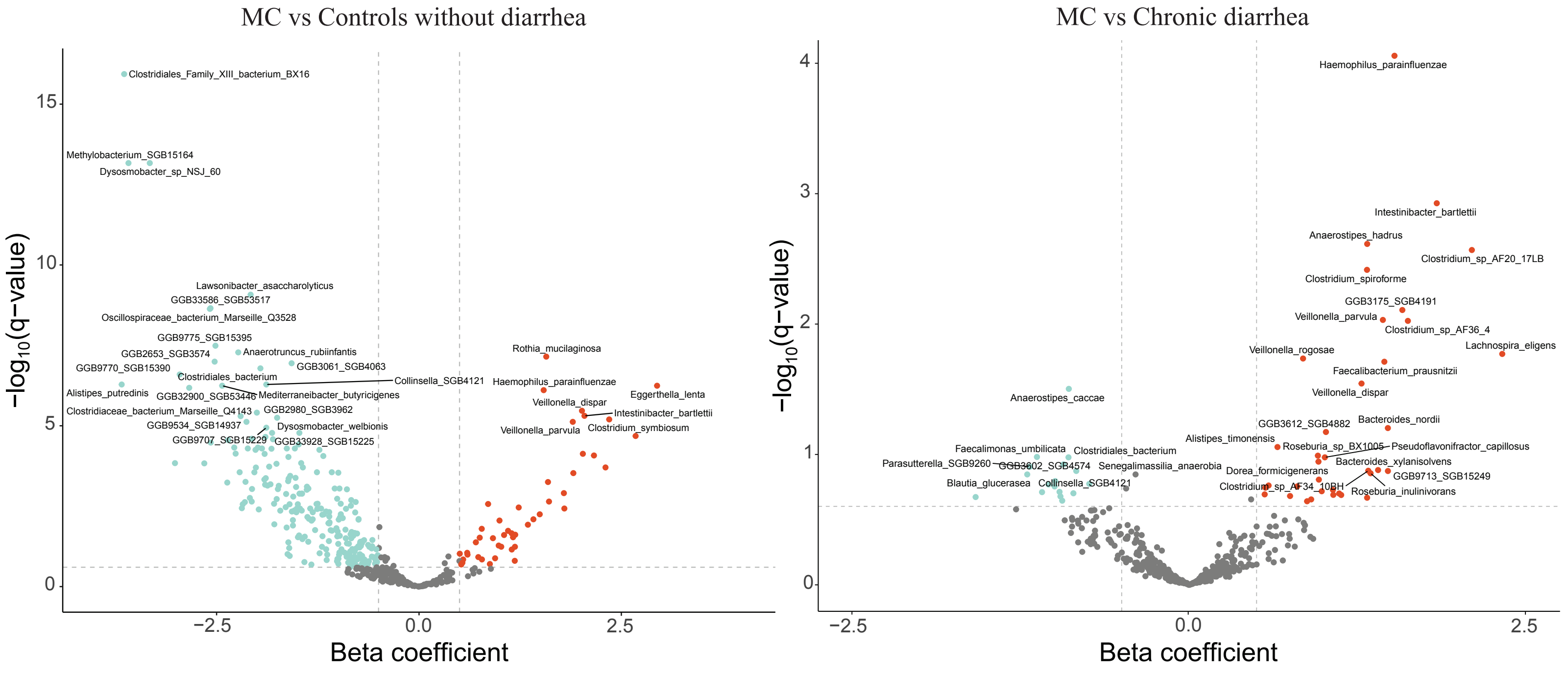


The volcano plots demonstrated alteration of species' relative abundance in MC compared to controls without diarrhea and chronic diarrhea controls. Coefficients and q-values were derived from MaAsLin using the aforementioned model. Red color indicated enriched relative abundance, turquoise color indicated depleted relative abundance, and gray color indicated no change in relative abundance.
