## Supplementary material for "Composition and Function of the Gut Microbiome in Microscopic Colitis": Fig S4

**S figure 4.** Comparisons of relative abundance of altered species according to MC subtypes.

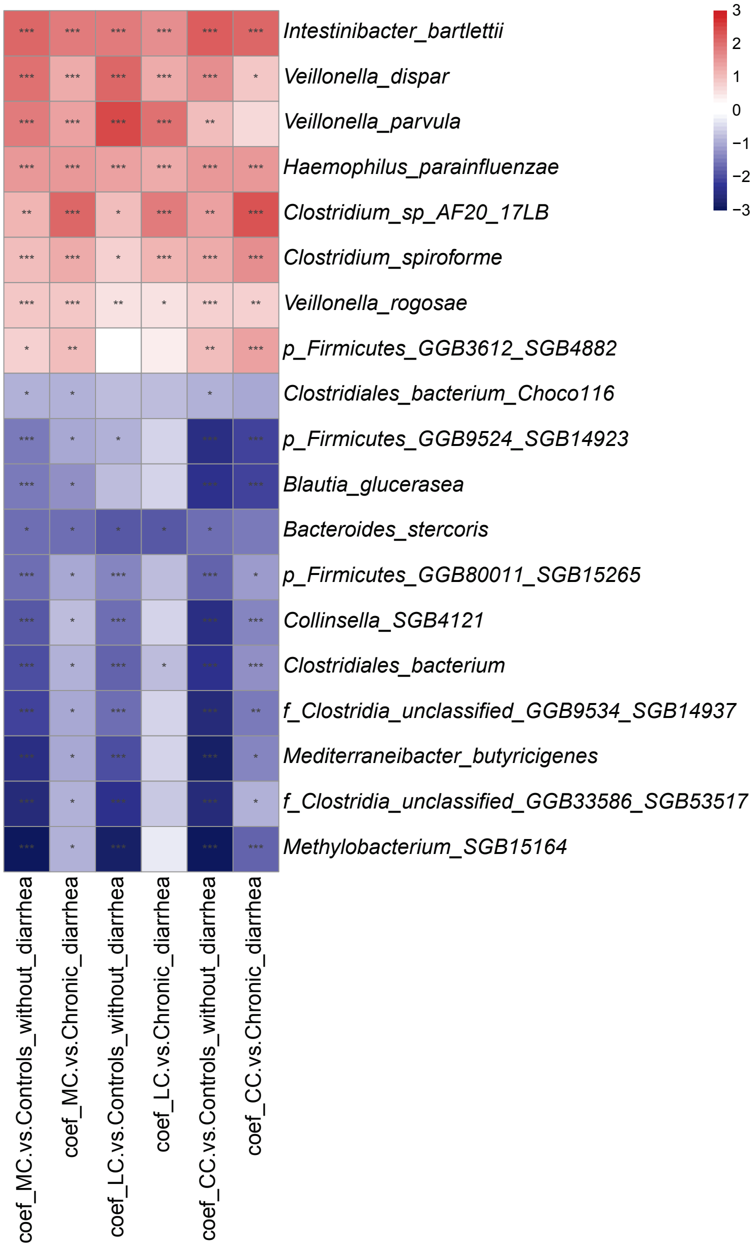


We compared the relative abundance of the 19 MC-altered species in LC and CC. Overall, MC-enriched species were also enriched in LC and CC, but MC-depleted species were only significantly depleted in CC but not in LC.
