## Supplementary material for "Composition and Function of the Gut Microbiome in Microscopic Colitis": Fig S5

**S figure 5.** Principal coordinate analysis based on the Bray-Curtis dissimilarity of metabolite composition.

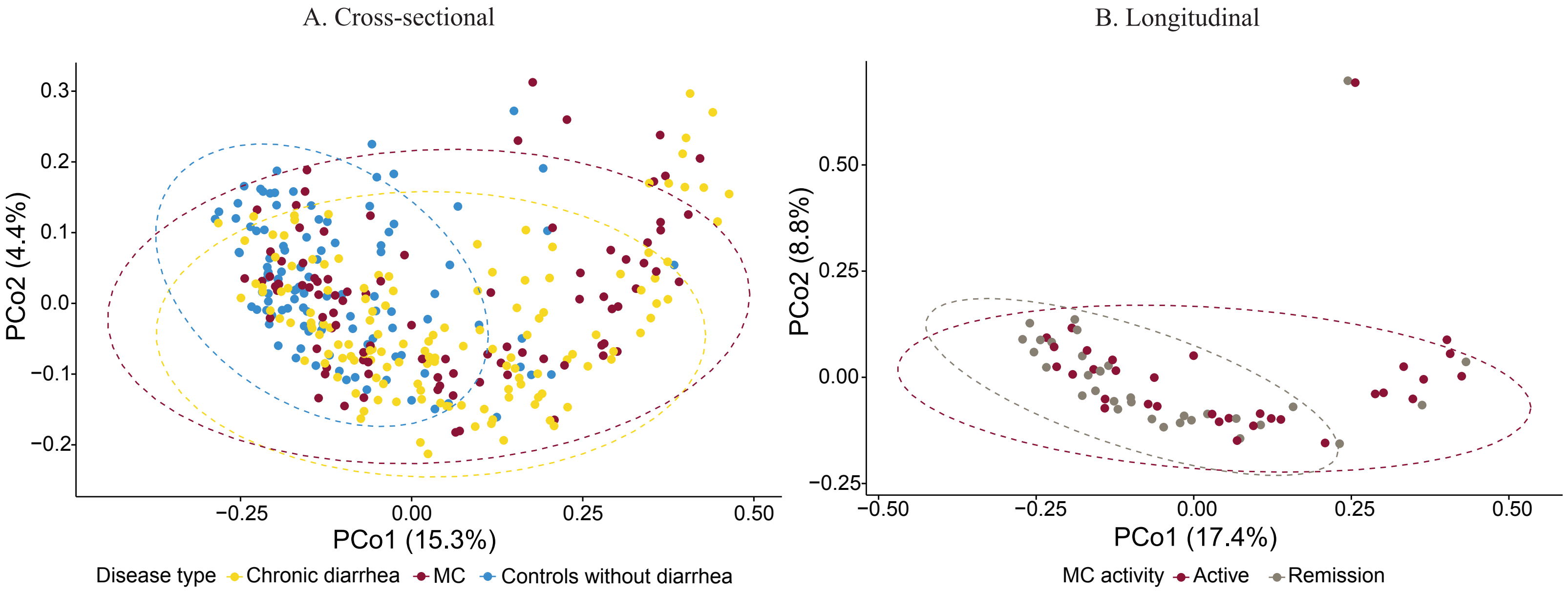


Among participants with both microbiome and metabolome data, principal coordinate analysis (PCoA) for metabolite composition showed relatively distinct distributions of MC vs controls without diarrhea (A) and active MC versus remission (B).
