## Supplementary material for "Composition and Function of the Gut Microbiome in Microscopic Colitis": Fig S6

**S figure 6.** Metabolite classes with altered abundance in MC compared to HC and CD in the cross-sectional cohort.

A. MC vs Controls without diarrhea

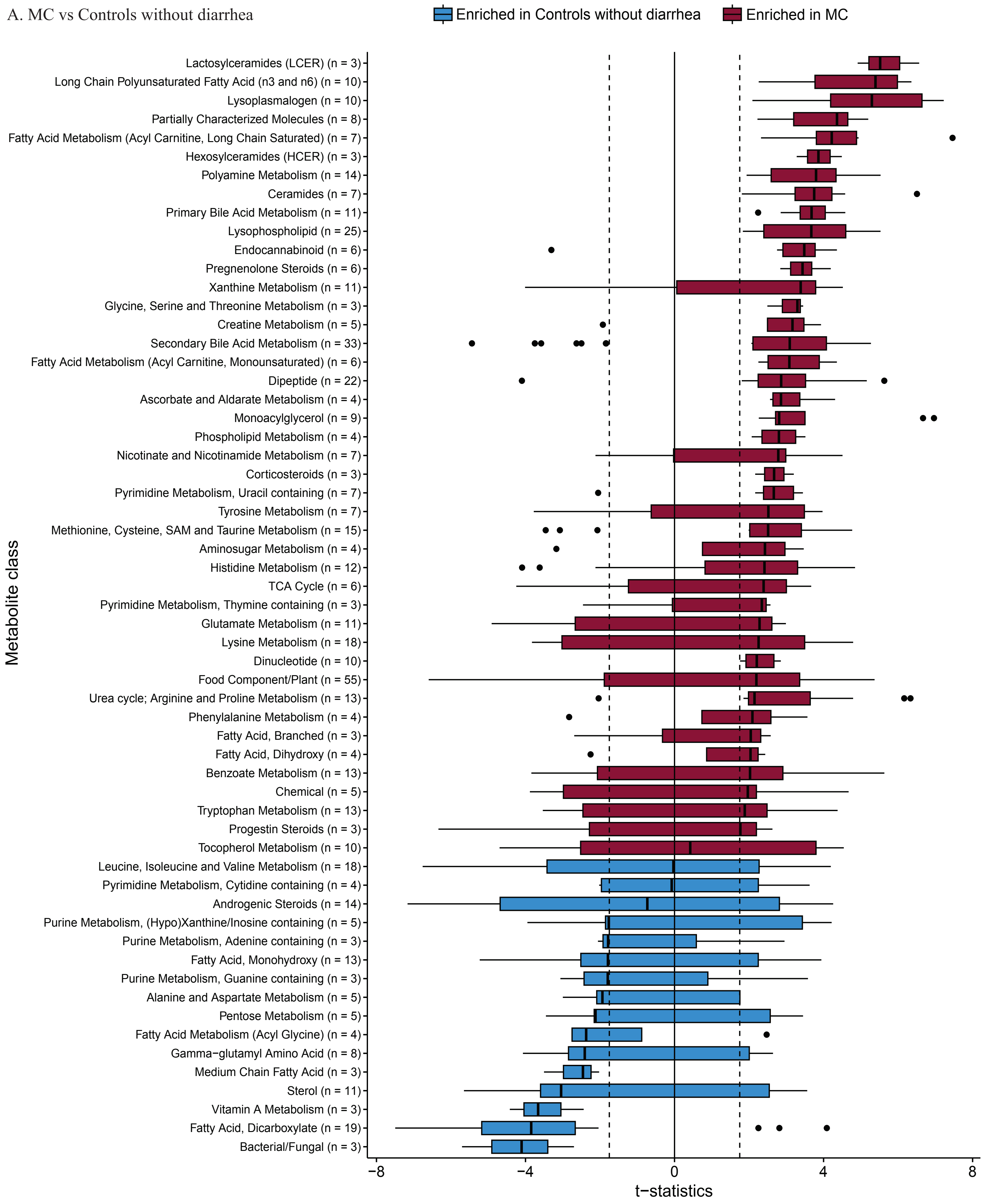
