## Supplementary material for "Composition and Function of the Gut Microbiome in Microscopic Colitis": Fig S7

**S figure 7.** Sphingolipid metabolism pathway [1, 2].

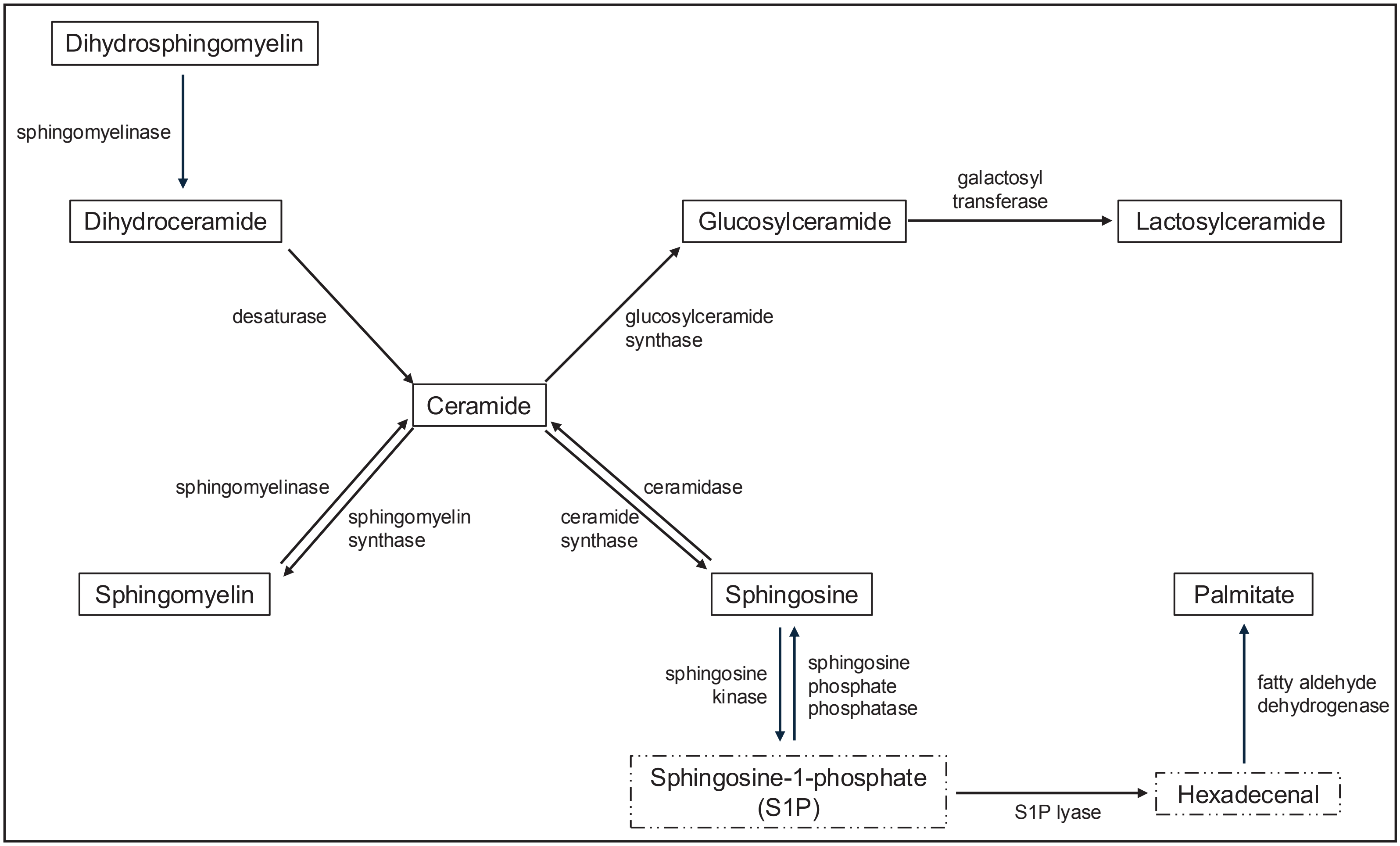


* Metabolites with dashed borders were not identified in our metabolomics analysis.
