## Supplementary material for "Composition and Function of the Gut Microbiome in Microscopic Colitis": Fig S8

**S figure 8.** Comparisons of relative abundance of altered metabolites according to disease type, MC subtypes, and activity.

A. LCER (top: cross-sectional; middle: LC; bottom: CC)

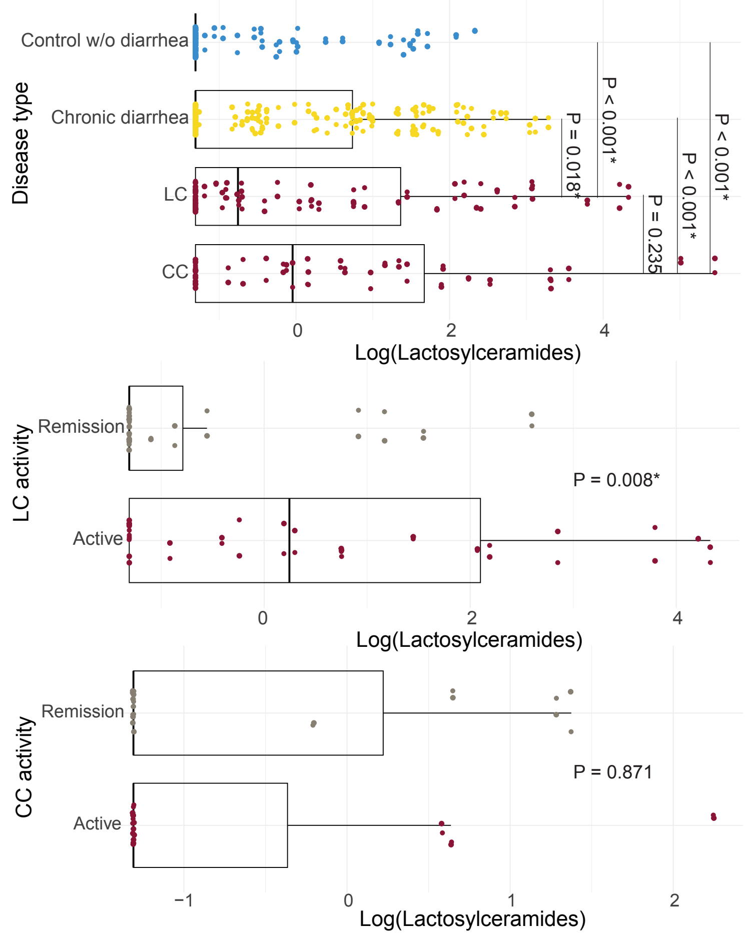


B. Ceramides (top: cross-sectional; middle: LC; bottom: CC)

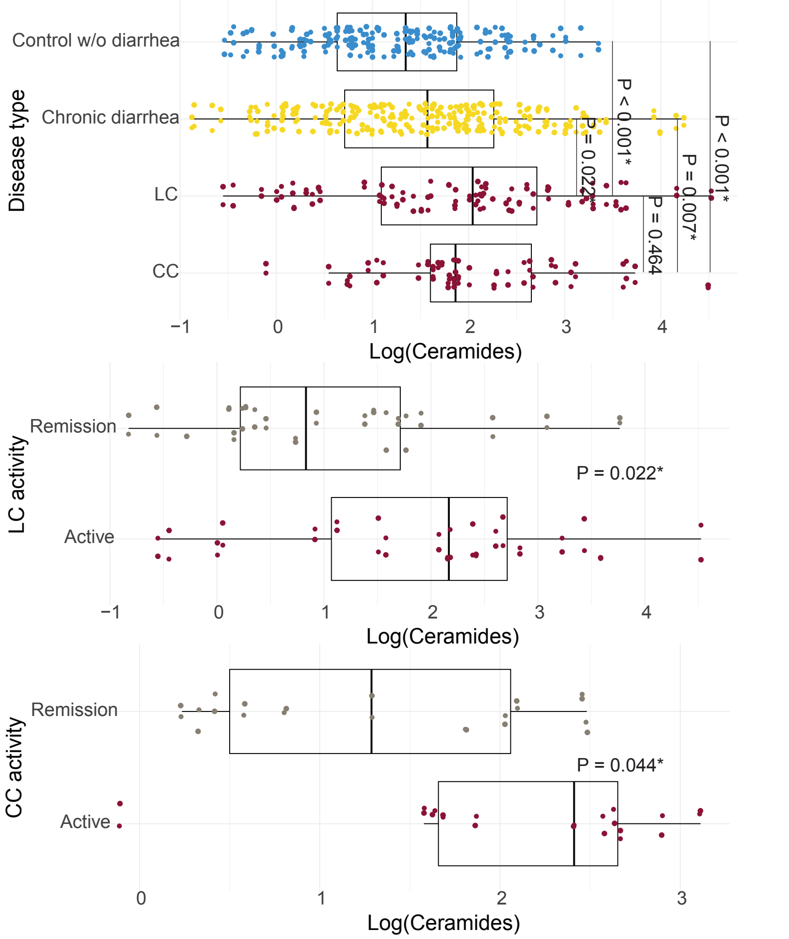


C. Lysophospholipids (top: cross-sectional; middle: LC; bottom: CC)

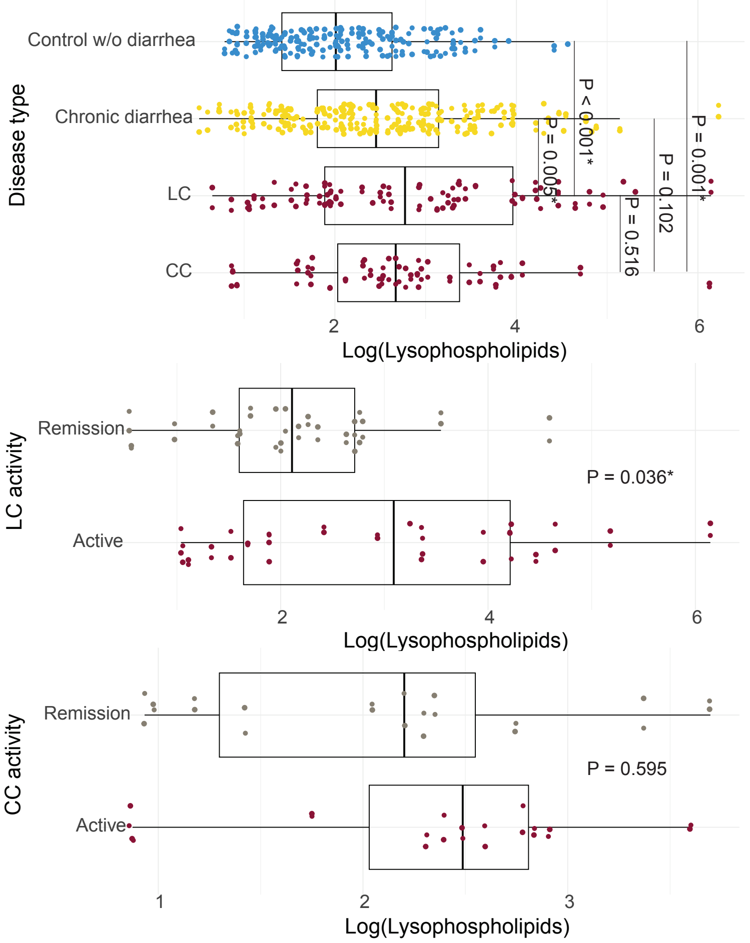


D. Lysoplasmalogens (top: cross-sectional; middle: LC; bottom: CC)

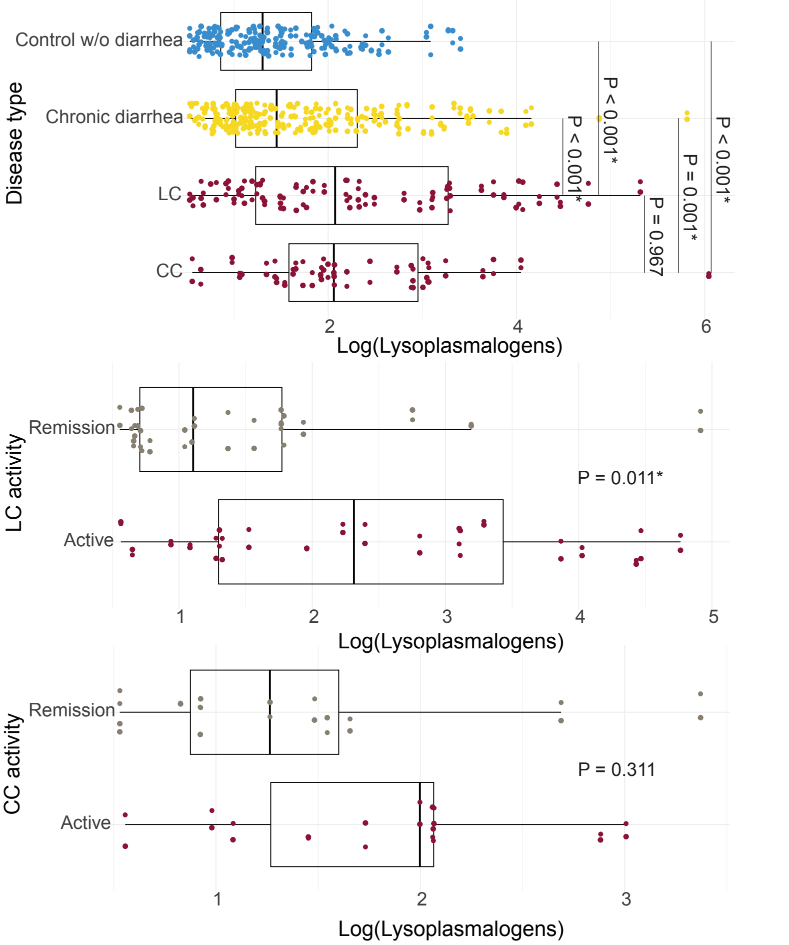

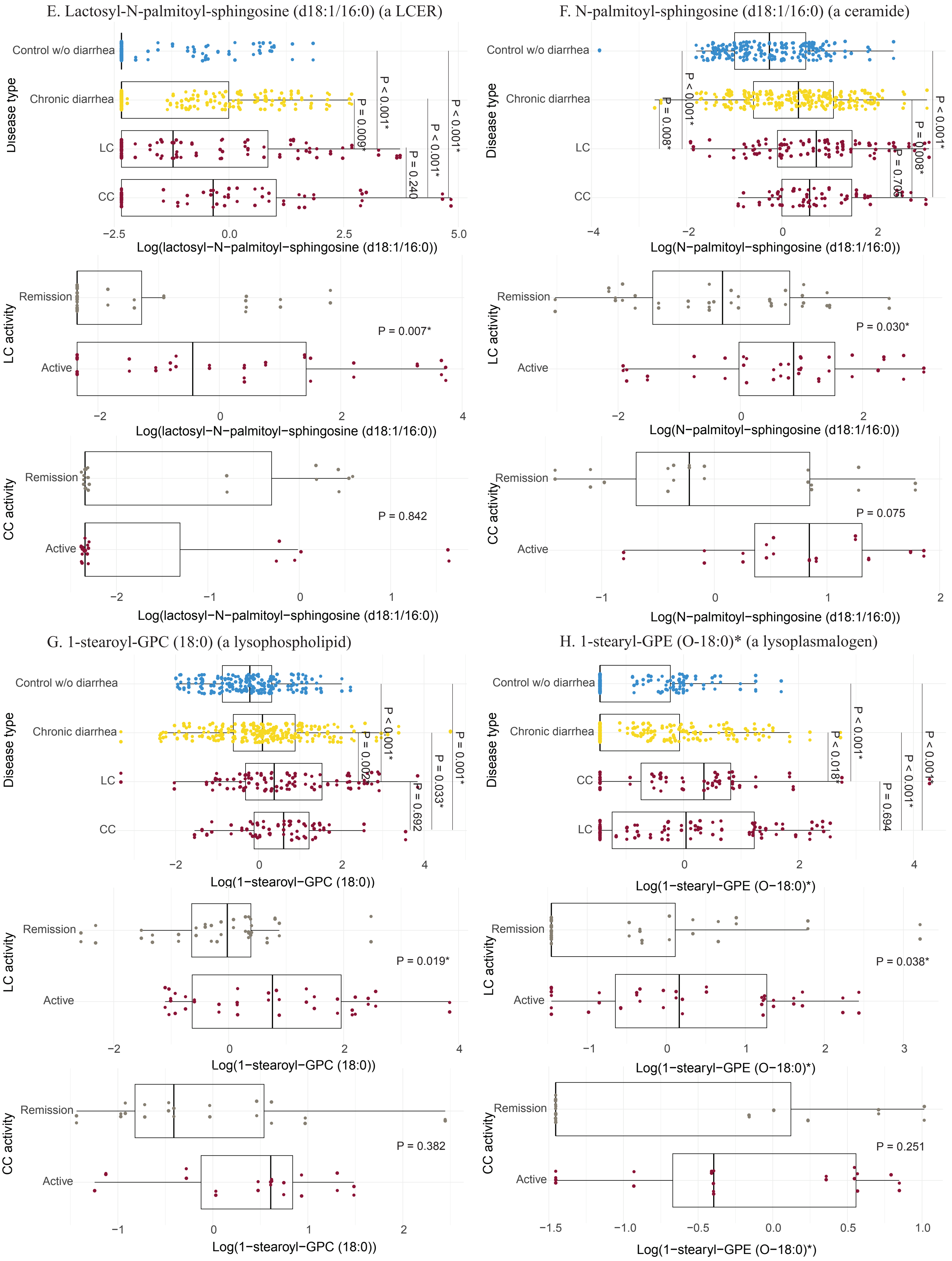

E. Lactosyl-N-palmitoyl-sphingosine (d18:1/16:0) (a LCER)
Control w/o diarrhea
Chronic diarrhea
LC
CC
Disease type
P < 0.001*
P = 0.009*
P < 0.001*
P = 0.240
P < 0.001*
−2.5
0.0
2.5
5.0
Log(lactosyl−N−palmitoyl−sphingosine (d18:1/16:0))
Remission
Active
LC activity
P = 0.007*
−2
0
2
4
Log(lactosyl−N−palmitoyl−sphingosine (d18:1/16:0))
Remission
Active
CC activity
P = 0.842
−2
−1
0
1
Log(lactosyl−N−palmitoyl−sphingosine (d18:1/16:0))
F. N-palmitoyl-sphingosine (d18:1/16:0) (a ceramide)
Control w/o diarrhea
Chronic diarrhea
LC
CC
Disease type
P = 0.008*
P < 0.001*
P = 0.706
P = 0.008*
P < 0.001*
−4
−2
0
2
Log(N−palmitoyl−sphingosine (d18:1/16:0))
Remission
Active
LC activity
P = 0.030*
−2
0
2
Log(N−palmitoyl−sphingosine (d18:1/16:0))
Remission
Active
CC activity
P = 0.075
−1
0
1
2
Log(N−palmitoyl−sphingosine (d18:1/16:0))
G. 1-stearoyl-GPC (18:0) (a lysophospholipid)
Control w/o diarrhea
Chronic diarrhea
LC
CC
Disease type
P = 0.002*
P < 0.001*
P = 0.692
P = 0.033*
P = 0.001*
−2
0
2
4
Log(1−stearoyl−GPC (18:0))
Remission
Active
LC activity
P = 0.019*
−2
0
2
4
Log(1−stearoyl−GPC (18:0))
Remission
Active
CC activity
P = 0.382
−1
0
1
2
Log(1−stearoyl−GPC (18:0))
H. 1-stearyl-GPE (O-18:0)* (a lysoplasmalogen)
Control w/o diarrhea
Chronic diarrhea
CC
LC
Disease type
P < 0.018*
P < 0.001*
P = 0.694
P < 0.001*
P < 0.001*
0
2
4
Log(1−stearyl−GPE (O−18:0)*)
Remission
Active
LC activity
P = 0.038*
−1
0
1
2
3
Log(1−stearyl−GPE (O−18:0)*)
Remission
Active
CC activity
P = 0.251
−1.5
−1.0
−0.5
0.0
0.5
1.0
Log(1−stearyl−GPE (O−18:0)*)
