## Supplementary material for "Composition and Function of the Gut Microbiome in Microscopic Colitis": Fig S9

### **S figure 9.** Correlations between microbiome composition and altered metabolites (complete heatmaps).

A. Correlations between microbial species and **sphingolipids**.

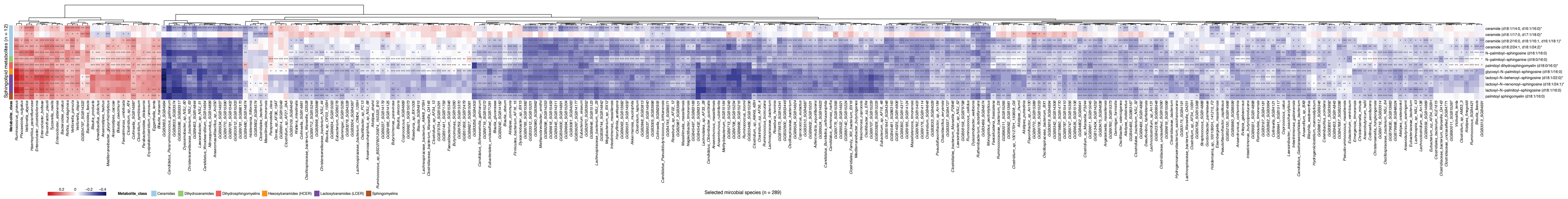


B. Correlations between microbial species and **lysophospholipids**.

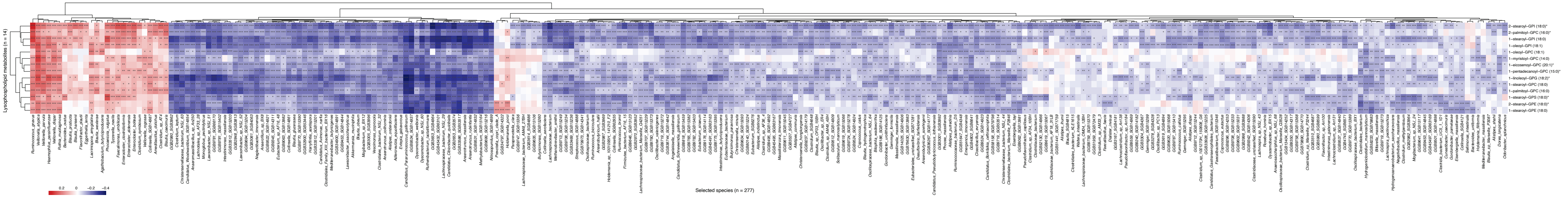


C. Correlations between microbial species and **lysophospholipids**.

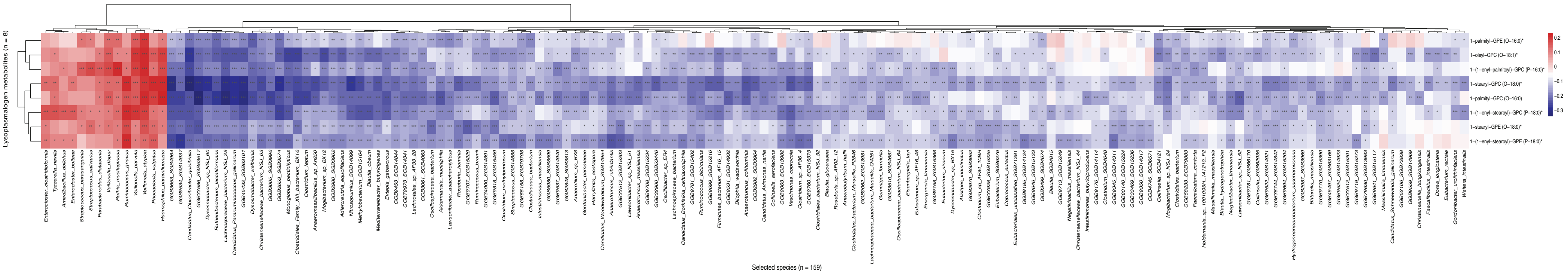


The complete heatmaps demonstrated all the microbial species highly correlated with (Spearman rho >0.15 or < -0.15) altered metabolites, including sphingolipids (A), lysophospholipids (B), lysoplasmalogens (C).
