## Supplementary material for "Composition and Function of the Gut Microbiome in Microscopic Colitis": Fig S10

**S figure 10.** Correlations between microbiome composition and altered metabolites (simplified heatmaps).

A. Correlations between highlighted microbial species and **sphingolipids**.

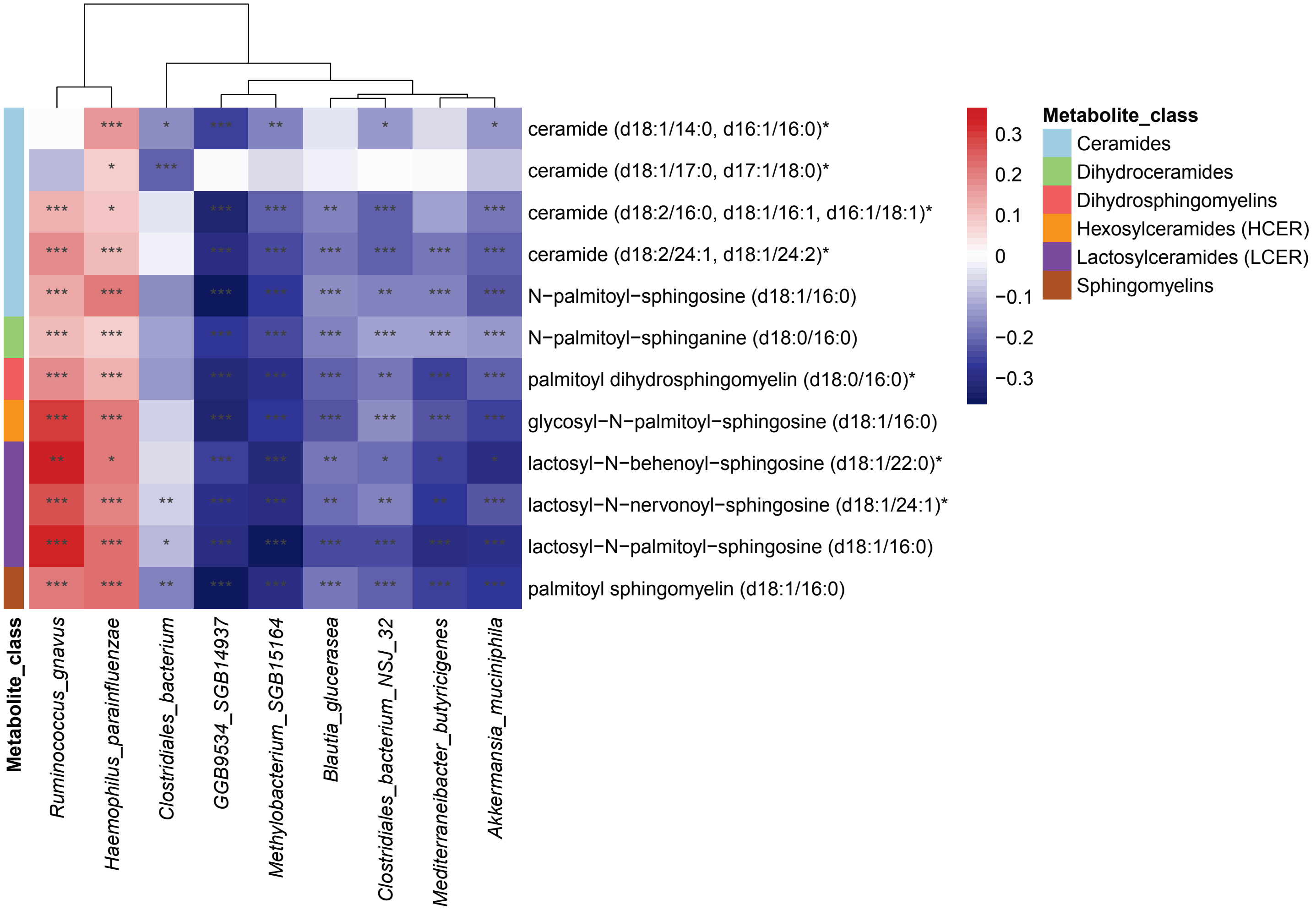

B. Correlations between highlighted microbial species and **lysophospholipids**.

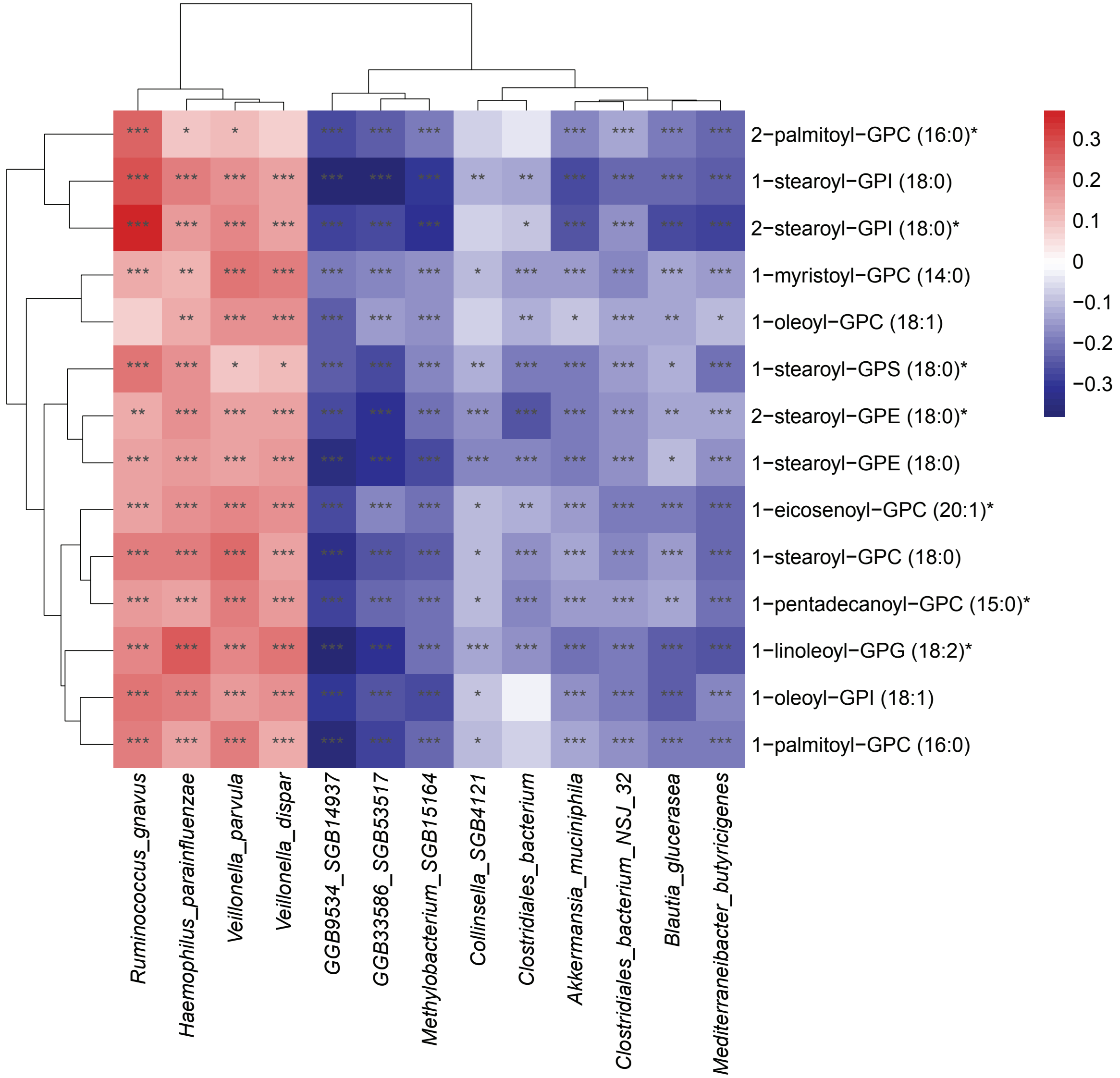

C. Correlations between highlighted microbial species and **lysoplasmalogens**.

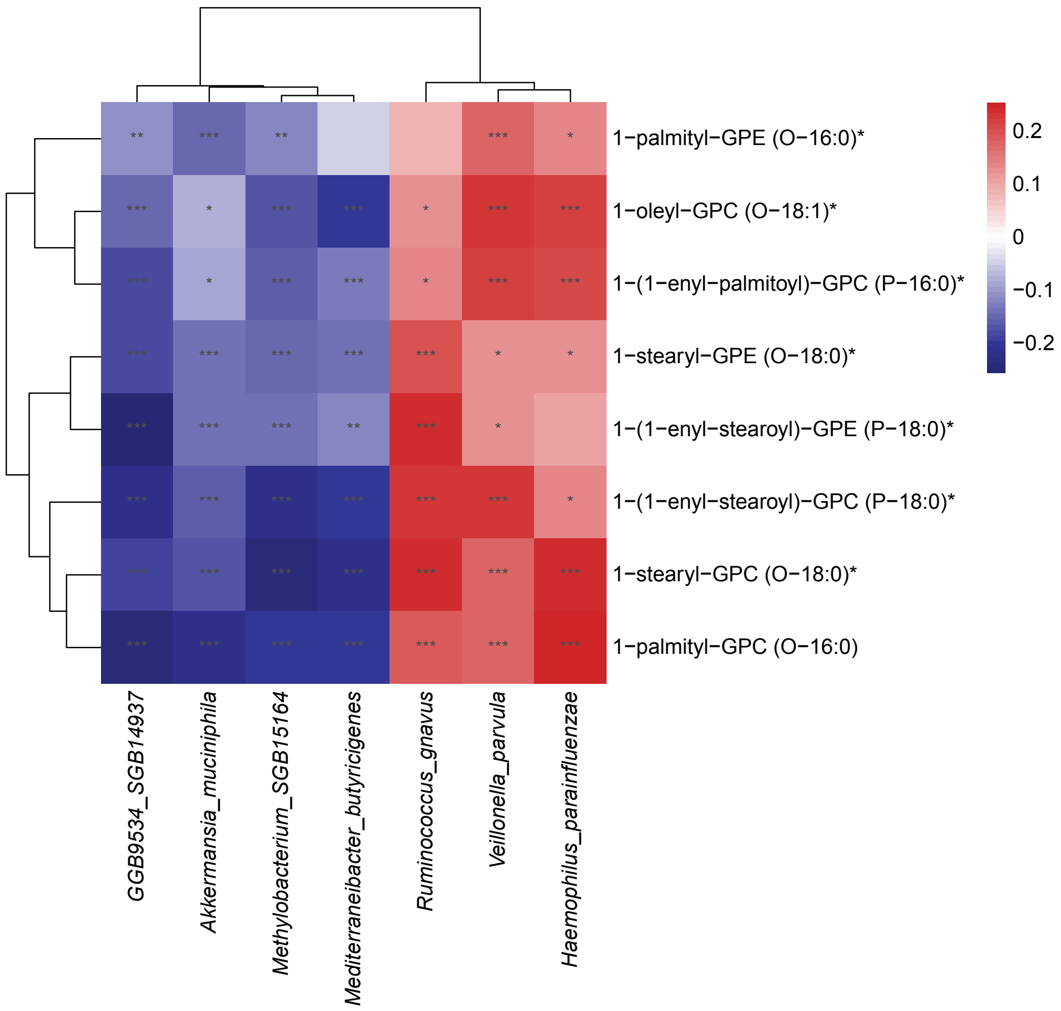


The simplified heatmaps showed selected species (those with altered abundance in MC and important MC species reported in literature) highly correlated with sphingolipids (A), lysophospholipids (B), lysoplasmalogens (C). Overall, pathologic species (*H. parainfluenzae*, *Veillonella spp.*, and *R. gnavus*) were positively correlated with the pro-inflammatory metabolites, while protective species (*Methylobacterium_SGB15164*, *B. glucerasea*, *M. butyricigenes*, and *A. muciniphila*) were negatively correlated with those metabolites.
