## Supplementary material for "Composition and Function of the Gut Microbiome in Microscopic Colitis": Fig S11

**S figure 11.** Scatter plots for species relative abundance and microbe-metabolite correlation coefficient.

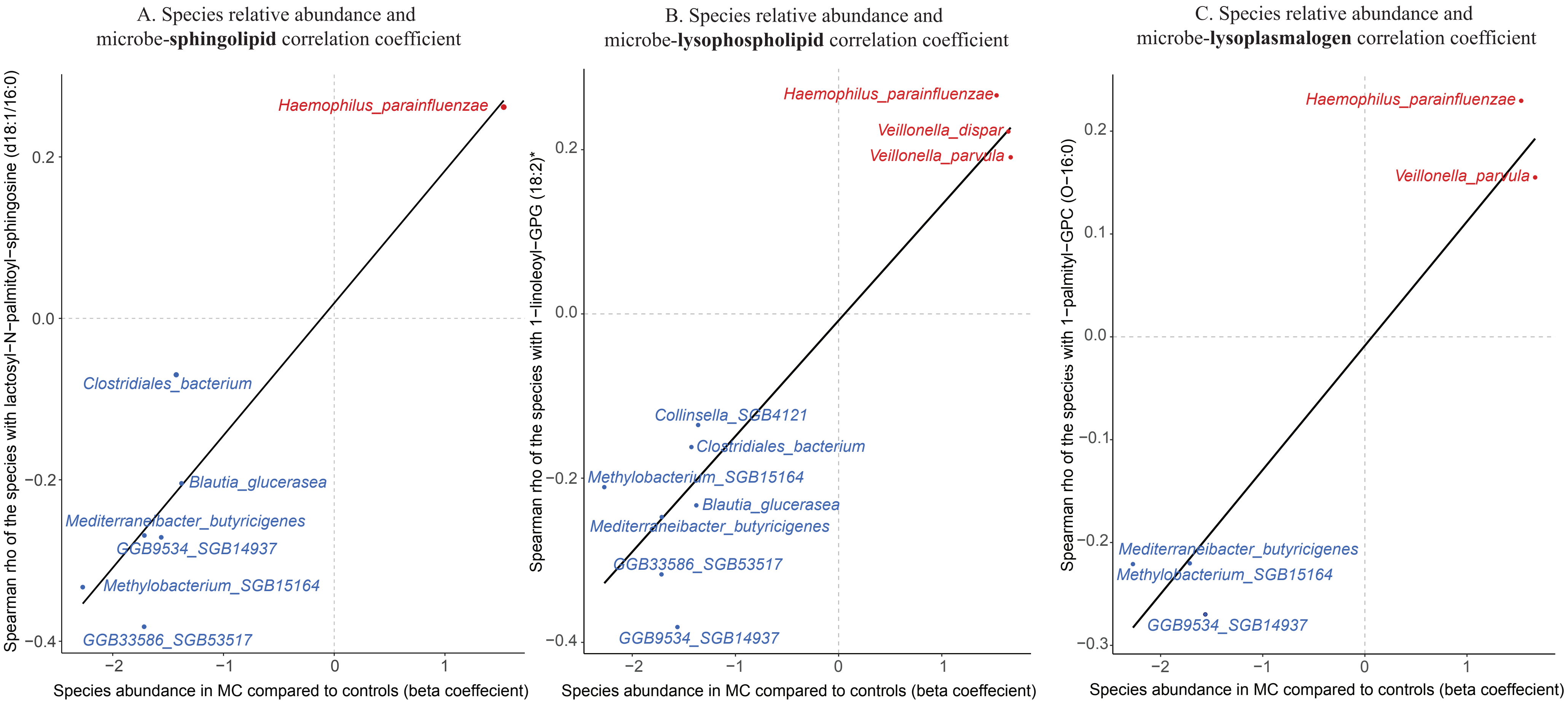


From **S figure 10**, we identified species strongly correlated with sphingolipids, lysophospholipids, and lysoplasmalogens, suggesting a functional correlation. Concordantly, some species also showed altered relative abundance in active MC versus controls and remission. We then plot these species onto the scatter plot. Each dot represented an individual species. The x-axis indicated the relative abundance of each species in MC, and the y-axis showed the microbe-metabolite (sphingolipids (A), lysophospholipids (B), and lysoplasmalogens (C)) correlation coefficient of the species.

Species enriched in MC were positively correlated with metabolite abundance, while those depleted in MC were negatively correlated with metabolite abundance. This concordance between differential abundance analysis and functional analysis suggested that pathologic species (such as *H. parainfluenzae* and *Veillonella spp.*) may produce pro-inflammatory metabolites in MC, or pro-inflammatory metabolites produced by MC gut may inhibit the growth of protective species (such as *Methylobacterium_SGB15164*, *M. butyricigenes*, and *B. glucerasea*).
