## Supplementary material for "Composition and Function of the Gut Microbiome in Microscopic Colitis": Fig S12

**S figure 12.** Correlations between microbial metabolic pathways and altered metabolites (complete heatmaps).

A. Correlations between microbial metabolic pathways and **sphingolipids**.

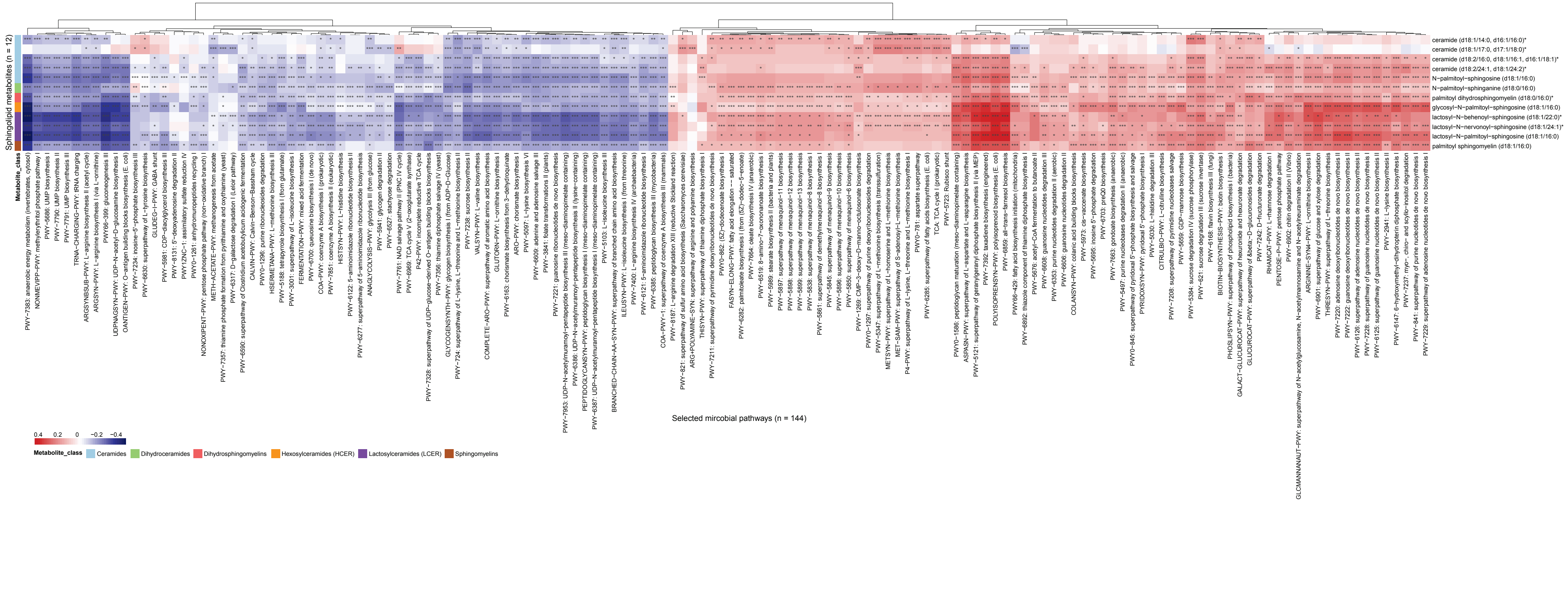

B. Correlations between microbial metabolic pathways and **lysophospholipids**.

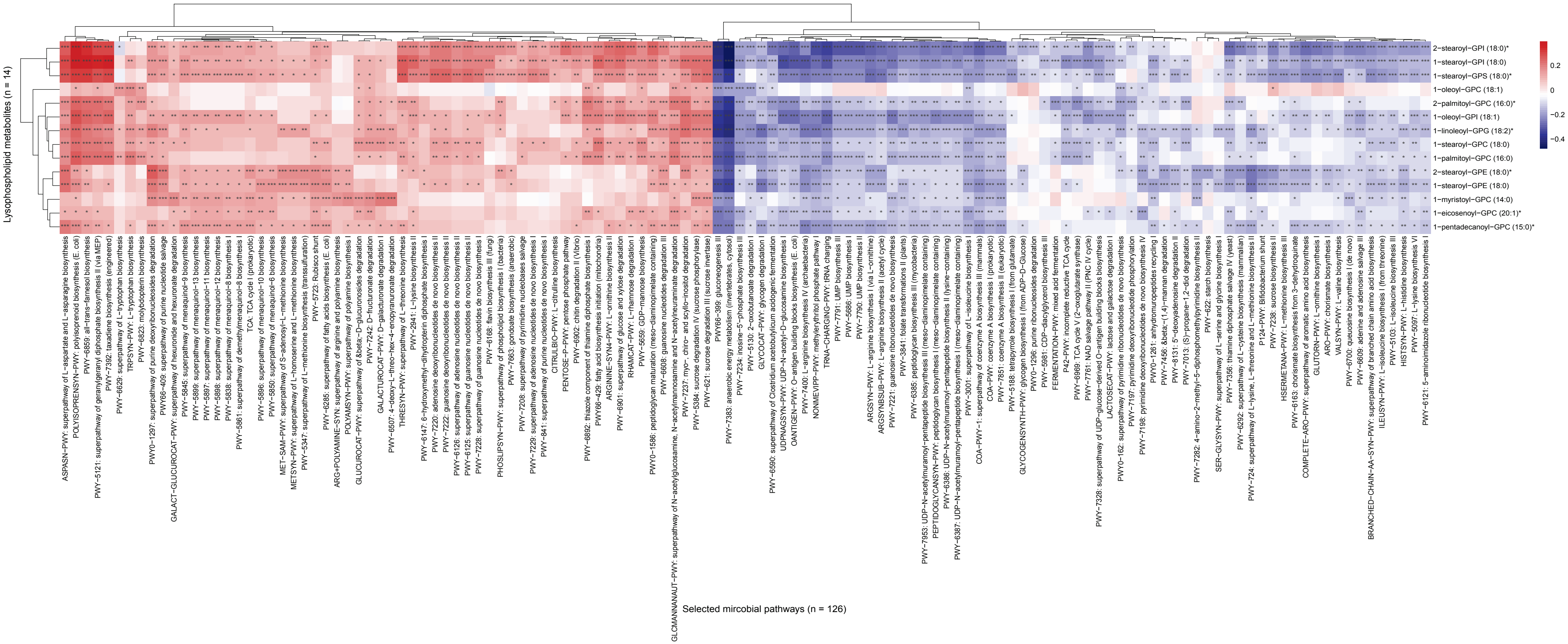

C. Correlations between microbial metabolic pathways and **lysoplasmalogens**.

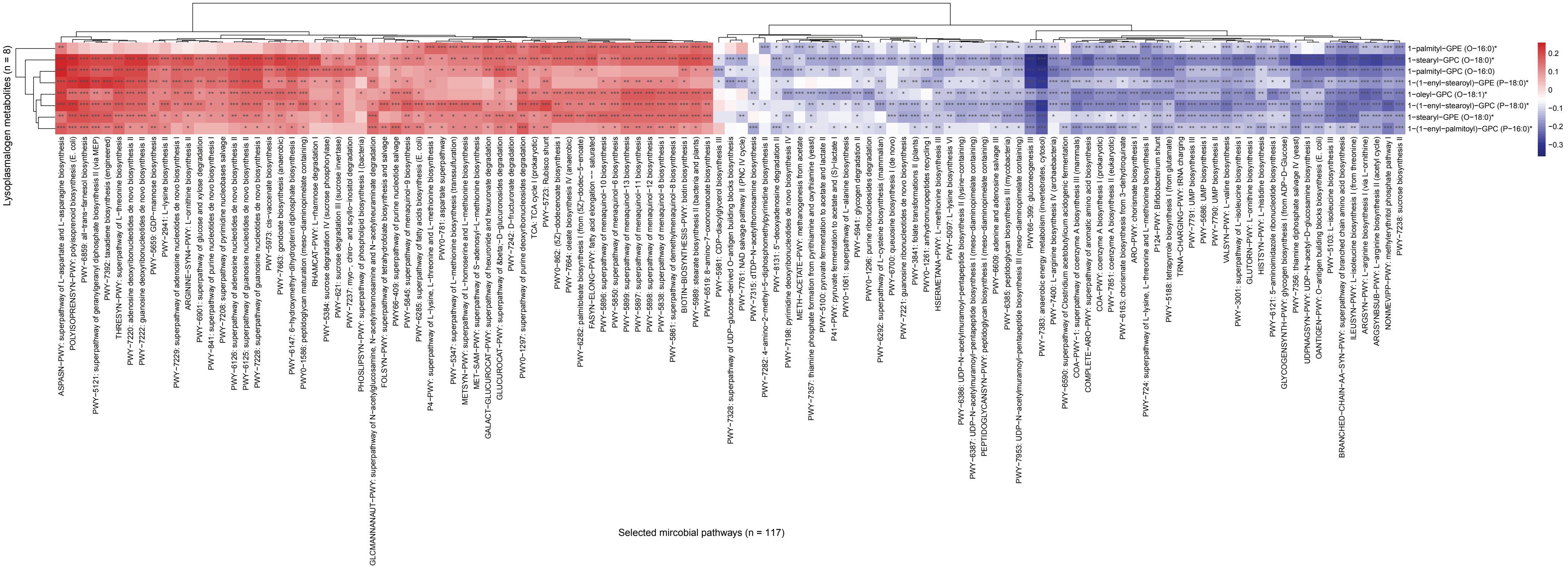


The complete heatmaps demonstrated all the microbial metabolic pathways highly correlated with (Spearman rho >0.15 or < -0.15) altered metabolites, including sphingolipids (A), lysophospholipids (B), lysoplasmalogens (C).
