## Supplementary material for "Composition and Function of the Gut Microbiome in Microscopic Colitis": Fig S13

**S figure 13.** Correlations between microbial metabolic pathways and altered metabolites (simplified heatmaps).

A. Correlations between highlighted microbial metabolic pathways and **sphingolipids**.

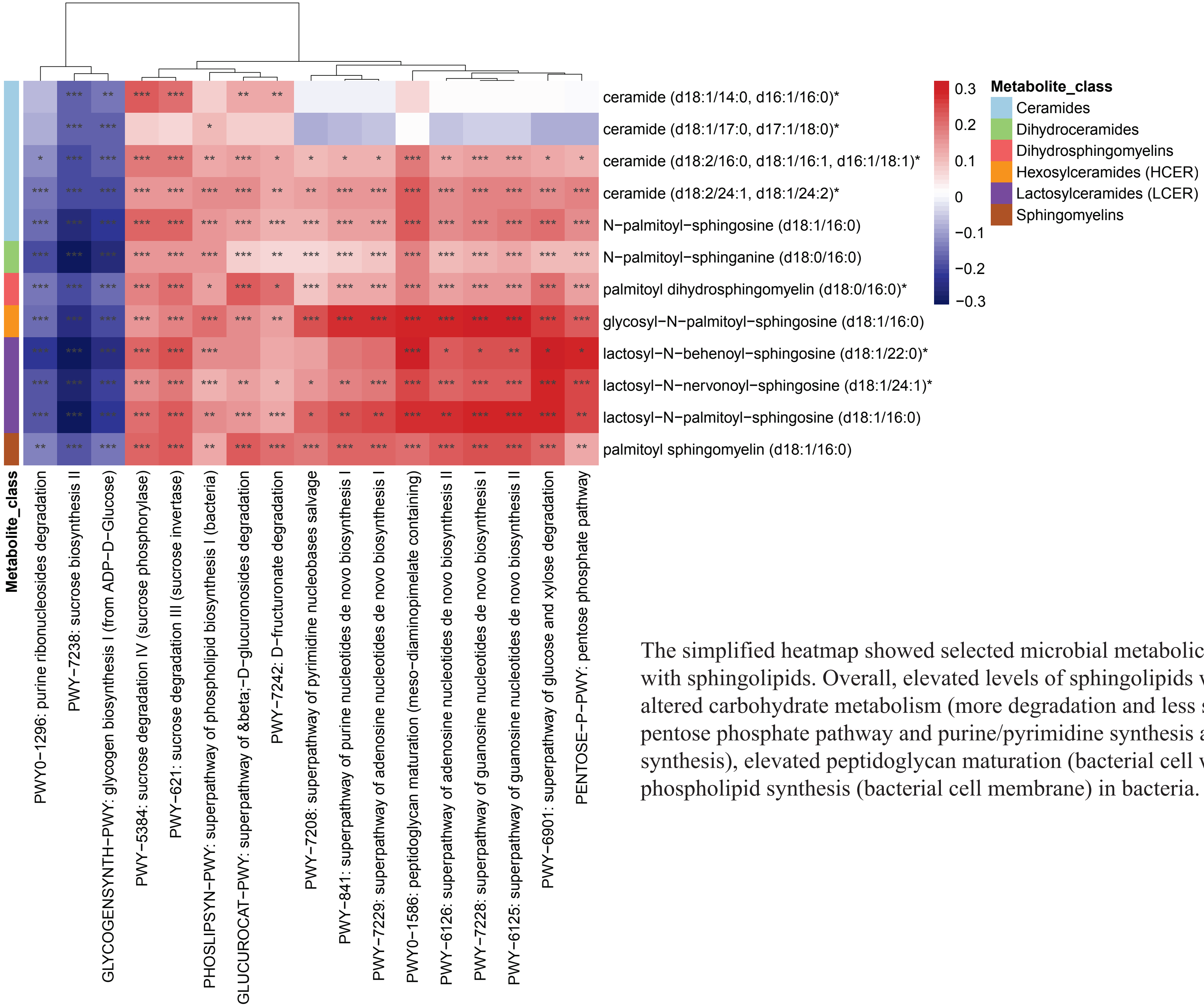


The simplified heatmap showed selected microbial metabolic pathways correlated with sphingolipids. Overall, elevated levels of sphingolipids were correlated with altered carbohydrate metabolism (more degradation and less synthesis), elevated pentose phosphate pathway and purine/pyrimidine synthesis and salvage (DNA/RNA synthesis), elevated peptidoglycan maturation (bacterial cell wall), and elevated phospholipid synthesis (bacterial cell membrane) in bacteria.

B. Correlations between highlighted microbial metabolic pathways and **lysophospholipids**.

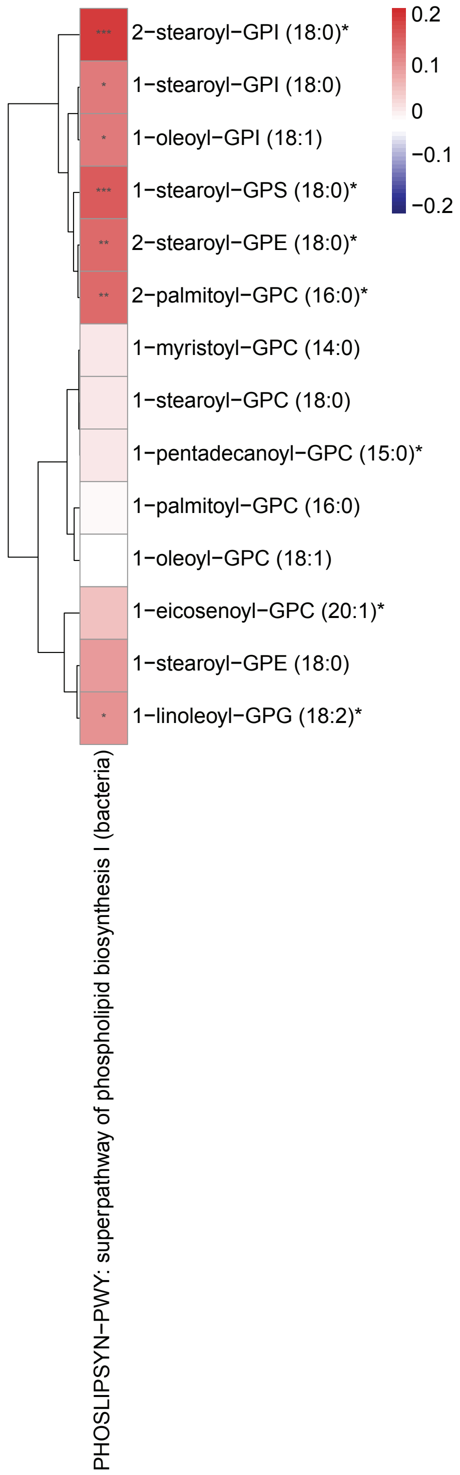


The simplified heatmap revealed selected microbial metabolic pathways correlated with lysophospholipids. Overall, elevated levels of lysophospholipids were correlated with upregulated phospholipid synthesis (the direct upstream metabolite of lysophospholipids) in bacteria, suggesting that the pro-inflammatory lysophospholipids may be produced by pathologic species and cause intraluminal inflammation in MC.

C. Correlations between highlighted microbial metabolic pathways and **lysoplasmalogens**.

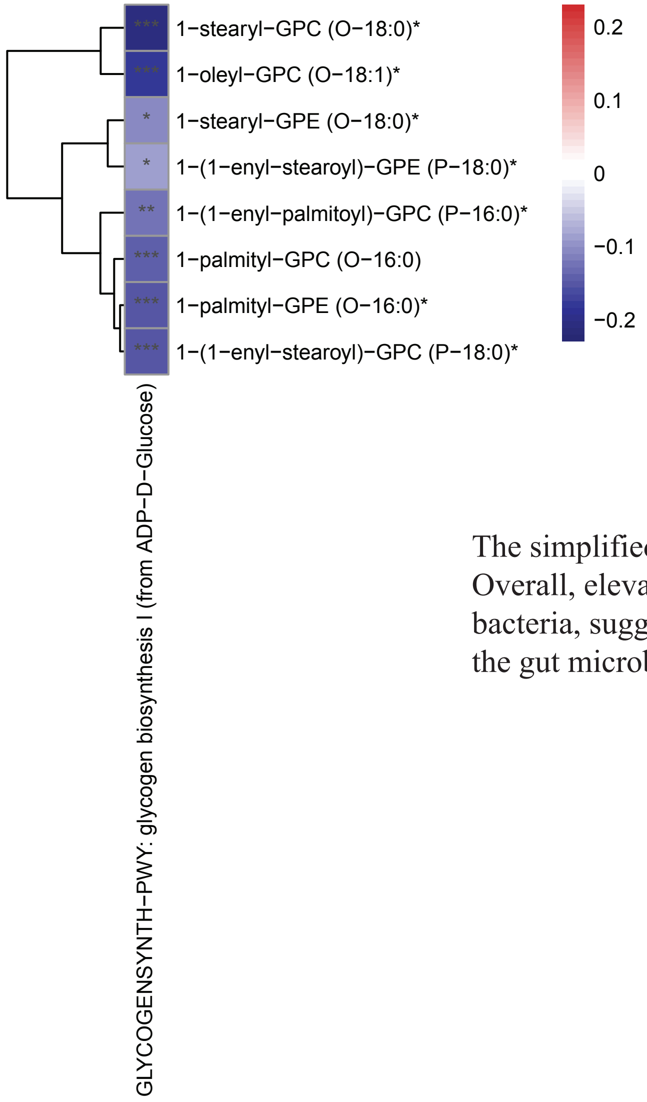


The simplified heatmap illustrated selected microbial metabolic pathways correlated with lysoplasmalogens. Overall, elevated levels of lysoplasmalogens were correlated with downregulated glycogen synthesis in bacteria, suggesting that the pro-inflammatory lysoplasmalogens in MC may cause environmental stress for the gut microbiome.
