## Supplementary material for "Composition and Function of the Gut Microbiome in Microscopic Colitis": Fig S14

**S figure 14.** Comparisons of relative abundance of metabolites related to S1P (A. upstream – sphingosine, B. downstream – palmitate) according to disease type and disease status.

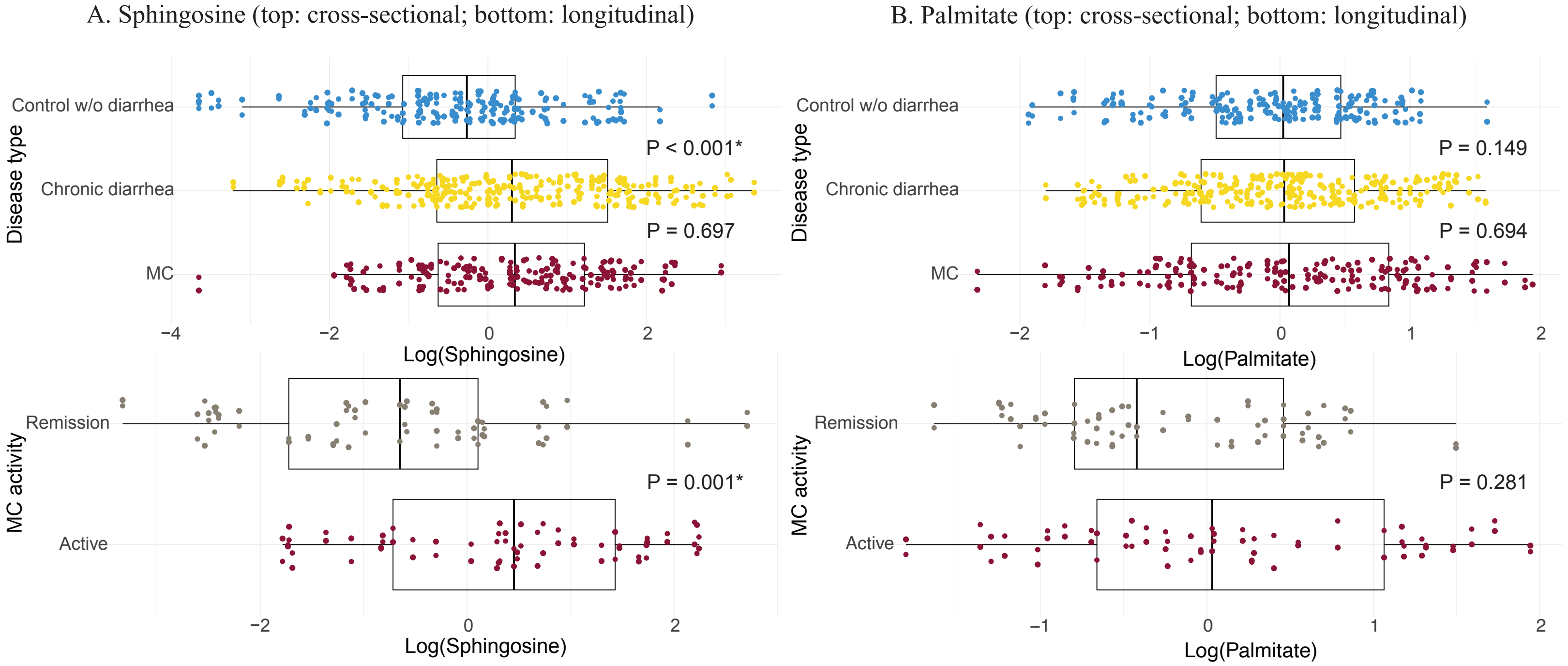


We observed higher sphingosine (upstream metabolite of S1P) levels in MC versus control without diarrhea and in active MC versus remission (A). However, palmitate (downstream metabolite of S1P) levels were similar in MC versus controls, and only a trend of enriched palmitate was observed in active MC versus remission (B).
